# Clinical validation of digital biomarkers and machine learning models for remote measurement of psoriasis and psoriatic arthritis

**DOI:** 10.1101/2022.04.13.22273676

**Authors:** Dan E. Webster, Rebecca H. Haberman, Lourdes Maria Perez Chada, Meghasyam Tummalacherla, Aryton Tediarjo, Vijay Yadav, Elias Chaibub Neto, Woody MacDuffie, Michael DePhillips, Eric Sieg, Sydney Catron, Carly Grant, Wynona Francis, Marina Nguyen, Muibat Yussuff, Rochelle L. Castillo, Di Yan, Andrea L. Neimann, Soumya M. Reddy, Alexis Ogdie, Athanassios Kolivras, Michael R. Kellen, Lara M. Mangravite, Solveig K. Sieberts, Larsson Omberg, Joseph F. Merola, Jose U. Scher

## Abstract

**Background:** Psoriasis and psoriatic arthritis are common immune-mediated inflammatory conditions that primarily affect the skin, joints and entheses and can lead to significant disability and worsening quality of life. Although early recognition and treatment can prevent the development of permanent damage, psoriatic disease remains underdiagnosed and undertreated due in part to the disparity between disease prevalence and relative lack of access to clinical specialists in dermatology and rheumatology. Remote patient self-assessment aided by smartphone sensor technology may be able to address these gaps in care, however, these innovative disease measurements require robust clinical validation.

**Methods:** We developed smartphone-based assessments, collectively named the *Psorcast* suite, that can be self-administered to measure cutaneous and musculoskeletal signs and symptoms of psoriatic disease. The image and motion sensor data collected by these assessments was processed to generate digital biomarkers or machine learning models to detect psoriatic disease phenotypes. To evaluate these digital endpoints, a cross-sectional, in-clinic validation study was performed with 92 participants across two specialized academic sites consisting of healthy controls and participants diagnosed with psoriasis and/or psoriatic arthritis.

**Findings:** In the domain of skin disease, digital patient assessment of percent body surface area (BSA) affected with psoriasis demonstrated very strong concordance (CCC = 0·94, [95%CI = 0·91–0·96]) with physician-assessed BSA. Patient-captured psoriatic plaque photos were remotely assessed by physicians and compared to in-clinic Physician Global Assessment parameters for the same plaque with fair to moderate concordance (CCC_erythema_=0·72 [0·59–0·85]; CCC_induration_=0·72 [0·62–0·82]; CCC_scaling_=0·60 [0·48–0·72]). Arm range of motion was measured by the *Digital Jar Open* assessment to classify physician-assessed upper extremity involvement with joint tenderness or enthesitis, demonstrating an AUROC = 0·68 (0·47–0·85). Patient-captured hand photos were processed with object detection and deep learning models to classify clinically-diagnosed nail psoriasis with an accuracy of 0·76, which is on par with remote physician rating of nail images (avg. accuracy = 0·63) with model performance maintaining accuracy when raters were too unsure or image quality was too poor for a remote assessment.

**Interpretation:** The *Psorcast* digital assessments, performed by patient self-measurement, achieve significant clinical validity when compared to in-person physical exams. These assessments should be considered appropriately validated for self-monitoring and exploratory research applications, particularly those that require frequent, remote disease measurements. However, further validation in larger cohorts will be necessary to demonstrate robustness and generalizability across populations for use in evidence-based medicine or clinical trial settings. The smartphone software and analysis pipelines from the *Psorcast* suite are open source and available to the scientific community.

**Funding:** This work is funded by the *Psorcast* Digital Biomarker Consortium consisting of Sage Bionetworks, Psoriasis and Psoriatic Arthritis Centers for Multicenter Advancement Network (PPACMAN), Novartis, UCB, Pfizer, and Janssen Pharmaceuticals. J.U.S work was supported by the Snyder Family Foundation and the Riley Family Foundation.

**Research in context:** *Evidence before this study:* No systematic literature review was performed. Patient self-measurement with smartphone sensors has been shown to be clinically valid for assessing signs and symptoms such as tremor, gait, physical activity, or range of motion across multiple disease indications. While smartphone-based applications have been developed for digitally tracking psoriatic disease, they have largely focused on questionnaire-based patient reported outcomes.

*Added value of this study:* To our knowledge, *Psorcast* is the first application using ubiquitous smartphone sensor technology for patients to remotely measure their psoriatic disease phenotypes, including detection of nail psoriasis and a continuous variable outcome measure of joint tenderness and enthesitis based on range of motion. This study not only developed a suite of novel, smartphone sensor-based assessment that can be self-administered to measure cutaneous and musculoskeletal signs and symptoms, but provides clinical validation of these measures.

*Implications of all the available evidence:* The developed *Psorcas*t suite of measurements can serve as groundwork for patient-driven, remote measurement of psoriatic disease. The use and continued development of this technology opens up new possibilities for both clinical care and research endeavors on a large scale. *Psorcast* measurements are currently being validated for their ability to assess disease changes longitudinally, allowing for more frequent symptom monitoring in clinical trials, more granular insight into the time course of medication action, and possible identification of responders from non-responders to specific therapies.

## Introduction

Psoriasis is one of the most common inflammatory skin conditions, affecting 2-3% of the US population and an estimated 125 million people worldwide.^1–3^ Up to 30% of individuals with psoriasis will develop psoriatic arthritis (PsA),^4^ an immune-mediated inflammatory arthritis involving the peripheral joints, spine, and entheses that can lead to permanent and disabling articular destruction.^5,6^ Patients with psoriatic disease (PsD), the spectrum of syndromes encompassing psoriasis and PsA, have worse quality of life than the general population with values similar to cancer, diabetes, and heart failure. PsD is also associated with a large economic burden to the healthcare system in terms of psychosocial disability and productivity loss.^7–12^

Despite the significant advances in therapeutics over the last two decades, multiple gaps in care remain, most notably underdiagnosis and undertreatment. Many patients with PsA may go undiagnosed for years after joint symptom onset.^13–15^ Even with a better understanding of clinical management and the development of clear treatment recommendations by multiple organizations,^16^ up to 39% of patients with moderate to severe psoriasis remain untreated and up to half of patients with PsA do not achieve clinically meaningful improvement with available medications.^17,18^ Perhaps most importantly, patients with PsD are dissatisfied with their care^17^ due in part to lack of access to, and dedication capacity of, trained clinical specialists in dermatology and rheumatology.^19^

Patient self-administration of disease assessments may provide an opportunity to address these gaps in clinical care through more timely outcome metrics to determine response to therapy and early identification of PsA among patients with psoriasis. The COVID-19 pandemic has further underlined the importance and utility of technology for remote care of patients.^20,21^ Notably, smartphones have a capacity for high-quality imaging, measurement of functional movement with sensor-based assessments, and they are ubiquitously used in the patient population. Smartphone sensor-based measurements have demonstrated clinical validity for remote patient symptom measurement in Parkinson’s Disease^22^, multiple sclerosis^23^, rheumatoid arthritis,^24,25,26^, diabetes^27^ and many others,^28^ but no validated toolset exists for psoriasis and psoriatic arthritis.^29^

The objective of this work was to develop and clinically validate^30^ a suite of novel, smartphone sensor-based assessments that can be self-administered to measure cutaneous and musculoskeletal signs and symptoms of psoriatic disease. Towards this end, here we describe a cross-sectional study that investigated the validity of self-administered digital assessments relative to physician-assessed clinical endpoints for psoriasis and psoriatic arthritis.

## Methods

### Study recruitment and patient population

Adult patients with psoriasis (n=14), psoriatic arthritis (n=69), and healthy controls (n=9) were recruited from two academic centers, the New York University (NYU) Langone Psoriatic Arthritis Center and Brigham and Women’s Hospital (BWH), between June 5, 2019 to November 10, 2021. Diagnoses of psoriasis and PsA were confirmed by trained rheumatologists and/or dermatologists specializing in psoriatic disease. Patients with PsA fulfilled CASPAR criteria.^10^ All treatments were determined by their primary physicians and patients may or may not have been receiving treatment with topical and/or systemic therapy for their psoriatic disease. This study was approved by the Institutional Review Boards of both NYU and BWH.

### In-clinic evaluation and data collection

Patient demographics and clinical evaluation was captured using a HIPAA-compliant web form implemented on the Synapse platform.^31^ Participants were evaluated by a trained rheumatologist and/or dermatologist. Clinical assessments included a musculoskeletal exam for tender and swollen joints, enthesitis, and dactylitis as well as a cutaneous exam with estimated percent body surface area (%BSA), physician global assessment, area of psoriasis involvement, and nail exam. On the same day, patients completed the *Psorcast* digital measures on a provided iPhone.

### Digital Assessment Design, Usability Studies and Development

Digital assessments were designed and developed based on important aspects of the disease,^32,33^ combined impact to patients and physicians (appendix p 5), and feasibility of being translated to a biometric measurement using smartphone sensors (Table 1). The assessments included: (1) *Psoriasis Draw*, (2) *Psoriasis Area Photo*, (3) *Painful Joint Count*, (4) *Digital Jar Open*, (5) *Fingers Photo*, (6) *Toes Photo*, and (7) *30-Second Walk*. All assessments were developed for the iOS operating system, and a full description and user interface design of each assessment can be found in appendix pp 2-4. Foundational user research and usability studies were conducted with psoriasis and psoriatic arthritis patients. These led to the development of multiple user experience features within the digital assessments, notably: the inclusion of written, animated, and video instructions; titration for most accurate *Psoriasis Draw* stroke width; elimination of complex touch gestures due to functional impairment of the hands in the target population; and immediate return of visualized results from the digital assessment.

**Table 1.** Digital assessment validation approach and outcomes.

| Physical Exam Clinical Endpoint Comparator | Psoriatic Disease Domain | Digital Endpoint Comparator | Digital Assessment Description | Validation Outcome Statistics |
| --- | --- | --- | --- | --- |
| Physician BSA (%) | Skin | Patient digital BSA (%) | *Psoriasis Draw*: Touch-screen body map drawing capturing location, extent of involvement | CCC = 0.94 (0.91–0.96) |
| Physician PGA | Skin | Physician panel remote PGA of patient image | *Psoriasis Area Photo*: Smartphone imaging of patient-selected representative area of the body with psoriasis | CCC<sub>Erythema</sub> = 0.72 (0.59–0.85)<br>CCC<sub>Induration</sub> = 0.72 (0.62–0.82)<br>CCC<sub>Scaling</sub> = 0.60 (0.48–0.72) |
| Physician tender joint count | Peripheral Arthritis | Patient self-assessed tender joint count | *Painful Joint Count*: Tap-based painful joint selection on a body map | CCC = 0.33 (0.20–0.45) |
| Physician tender joint count and enthesitis assessment (upper extremities) | Peripheral Arthritis | Classification model detecting upper extremity involvement | *Digital Jar Open*: Gyroscope-based arm rotation measure with logistic regression model of upper extremity involvement | AUROC = 0.68 (0.47–0.85) |
| Physician nail assessment | Nail Psoriasis | Physician panel remote nail assessment of patient image | *Finger Photo*: Smartphone imaging of each hand with object detection and classification models to detect nail psoriasis | Accuracy = 0.63 (0.42–0.84)<br>Precision<sub>Nail PsO</sub> = 0.16 (0.08–0.24)<br>Recall<sub>Nail PsO</sub> = 0.56 (0.96–0.16) |
| Physician nail assessment | Nail Psoriasis | Classification model of nail psoriasis |  | Accuracy = 0.76 (0.75–0.76)<br>Precision<sub>Nail PsO</sub> = 0.93 (0.93–0.94)<br>Recall<sub>Nail PsO</sub> = 0.42 (0.42–0.43) |

### *Psoriasis Draw* digital body surface area (BSA) endpoint validation analysis

Percentage of the body surface area (BSA) affected by psoriasis was determined by dividing the number of pixels drawn on the body divided by the total number of pixels on the body. Each overall area (front-upper, back-upper, front-lower, back-lower) represents approximately 25% of the total area. Digital BSA was compared with an in-clinic physician BSA estimate using a Bland-Altman analysis with the Lin’s Concordance Correlation Coefficient (CCC) as the metric for validation.

### *Psoriasis Area Photo* digital physician global assessment (PGA) endpoint validation analysis

In the *Psoriasis Area Photo* task, patients first selected an area of the body (out of a total of 60 zones) that they determined contained a plaque that is representative of their current disease state. Patients told the physician the area they chose and the physician rated this plaque for severity levels (0-4) along the axes of erythema, induration, and scaling. A photo was then taken of the representative plaque by the patient with either the front- or rear-facing iPhone camera. Plaque photos were then remotely presented to a panel of physician raters (see ***remote image assessment*** section) In-clinic PGA ratings were compared to remote PGA ratings with concordance determined by Lin’s CCC.

### *Painful Joint Count* digital tender joint count endpoint validation analysis

Patients first selected one or more areas of the body (upper, lower, hands, feet) that contained painful joints. Within these areas, selectable joints included the following (left and right for each): shoulders, elbows, wrists, finger proximal interphalangeal (PIP) joints, finger distal interphalangeal joints, metacarpophalangeal joints, carpometacarpal joints, hips, knees, ankles, metatarsophalangeal, toe PIPs. Other joints typically included in clinician reported outcomes like the 66/68 joint count were excluded due to low levels of involvement in psoriasis populations and difficulty in participant recognition of more obscure joints. (e.g. including the “shoulder joint” instead of the glenohumeral and acromioclavicular joints). Patients were not asked to differentiate between tenderness and swelling. Physician-assessed tender joints were compared to patient-reported painful joints with Bland-Altman analysis.

### *Digital Jar Open* classification of upper extremity involvement (UEI) and PsA without UEI

For the *Digital Jar Open* assessment, overall rotation was used to build logistic regression classifiers for: (i) predicting the presence of UEI versus its absence, where the UEI represents clinically-assessed arm joint tenderness (wrist, elbow, shoulder) or enthesitis (lateral epicondyle); (ii) predicting PsA in the subset of individuals with known PsA diagnosis who did not present with UEI.

Classification performance was evaluated using the area under the receiver operating characteristic curve (AUROC), and area under the precision-recall curve (AUPRC), using the PRROC R package.^34^ The classifiers were trained and evaluated on 1000 distinct random splits of the data into training and test sets, with half of the data used for training and half for testing. Because the label classes were unbalanced and a small percentage of participants contributed a reduced number of longitudinal records, we performed stratified random splits at the subject level where: (i) the proportion of positive and negative labels is preserved across the training and test sets; and (ii) the data of each subject is either assigned to the training set or to the test set, but never to both. Subject-wise random splits of data sets containing repeated measurements on the subjects were employed to avoid artifacts due to identity confounding,^35,36^ where the inadvertent inclusion of records of the same participant in both the training and test set leads to artificially over-optimistic classification performance.)

In order to evaluate whether the classification performance was better than a random guess we also built classifiers using shuffled labels, which provide information about the range of AUROC and AUPRC scores we would expect to see by chance for our test set size. Furthermore, for the AUROC metric we also report the 95% confidence interval using the bootstrap method implemented in the ci.auc function of the pROC R package.^37^

### Confounding adjustment analyses

We investigated whether age, gender, and study site were confounding the predictions from our UEI and PsA without UEI classifiers. A detailed description of our approaches to assess and adjust for observed confounders is presented in appendix pp 9-26.

### *30-Second Walk* lower extremity involvement

Patients were asked to walk for 30 seconds with the smartphone in their pocket or waistband. Gait features from motion sensor data were extracted with PDKit^38^ and compared between lower extremity involvement (physician-assessed tender joints in the hip, knee, or ankles, or enthesitis in the achilles tendons or medial femoral condyles).

### Remote image assessment

Images from the study were loaded into the web-based Synapse platform using a custom R Shiny application, here termed *mHealthAnnotator, which* is described in more detail at the following: https://sage-bionetworks.github.io/mhealthannotator/. *mHealthAnnotator* presented physician raters with the photo and appropriate action button for scoring corresponding images. For plaque photos, the plaque qualities of erythema, induration, and scaling were presented on a 0-4 scale, with an additional option for ‘cannot tell’. Cropped, single-nail images were presented in the annotation portal with options for nail psoriasis present/absent, as well as ‘unsure’ or ‘cannot tell’. Nails that could not be captured from the image processing due to poor quality or digit positioning were excluded from this analysis. All thumb images were excluded for the latter reason.

### Nail object detection and nail psoriasis classification models

A transfer learning approach was employed in our workflow to process hand images for the classification of nail psoriasis. To perform nail object detection, we fine-tuned a pre-trained “Detectron2^39^ Model (Faster RCNN R101 FPN 3x)” on our custom nails dataset consisting of more than 5,000 individual nails segmented from open source images. For a psoriasis classifier, we used VGG16^40^ pretrained weights (imagenet) to train a binary classifier on the segmented nail images generated by the object detection model. For more elaborate discussion about our methods, see appendix pp 27-32.

### Hand image processing and Fitzpatrick skin tone estimation

Hand images were captured using the smartphone rear-facing camera and then processed using the Mediapipe framework (https://mediapipe.dev) to generate hand landmarks in the palm and joints of the fingers. Using these landmarks, we isolated a triangular region on the back of the hand and converted this RGB image into the ITA color space and then summarized the pixel values using a median. This summarized metric is then converted into a skin tone based on thresholds for the Fitzpatrick skin tone scale.^41^ OpenCV (https://opencv.org) was used for all image and pixel manipulations. Further detail on the image processing of the hands can be found in supplemental methods (appendix pp 33-37).

## Results

To investigate the clinical validity of self-administered *Psorcast* digital measurements, 92 participants were enrolled between two specialized academic sites. This cohort was balanced across age deciles and sex, with study participants exhibiting representative levels of cutaneous, musculoskeletal, and extra-articular signs and symptoms of disease^42^ (Fig. 1A,B; appendix pp 7-8). Using the participant’s self-reported plaque location from the *Psoriasis Draw* assessment, a high-resolution heatmap of plaque location and prevalence for the entire cohort was created. This heatmap demonstrated enrichment for known hotspots including the scalp, ears, elbows, lower back, and genital area (Fig. 1C). The cohort comprises a spectrum of pigmentation levels as measured by a novel photo-based digital estimation of skin tone adapted from the Fitzpatrick scale.^43,44^ (Fig. 1D).

**Figure 1.**
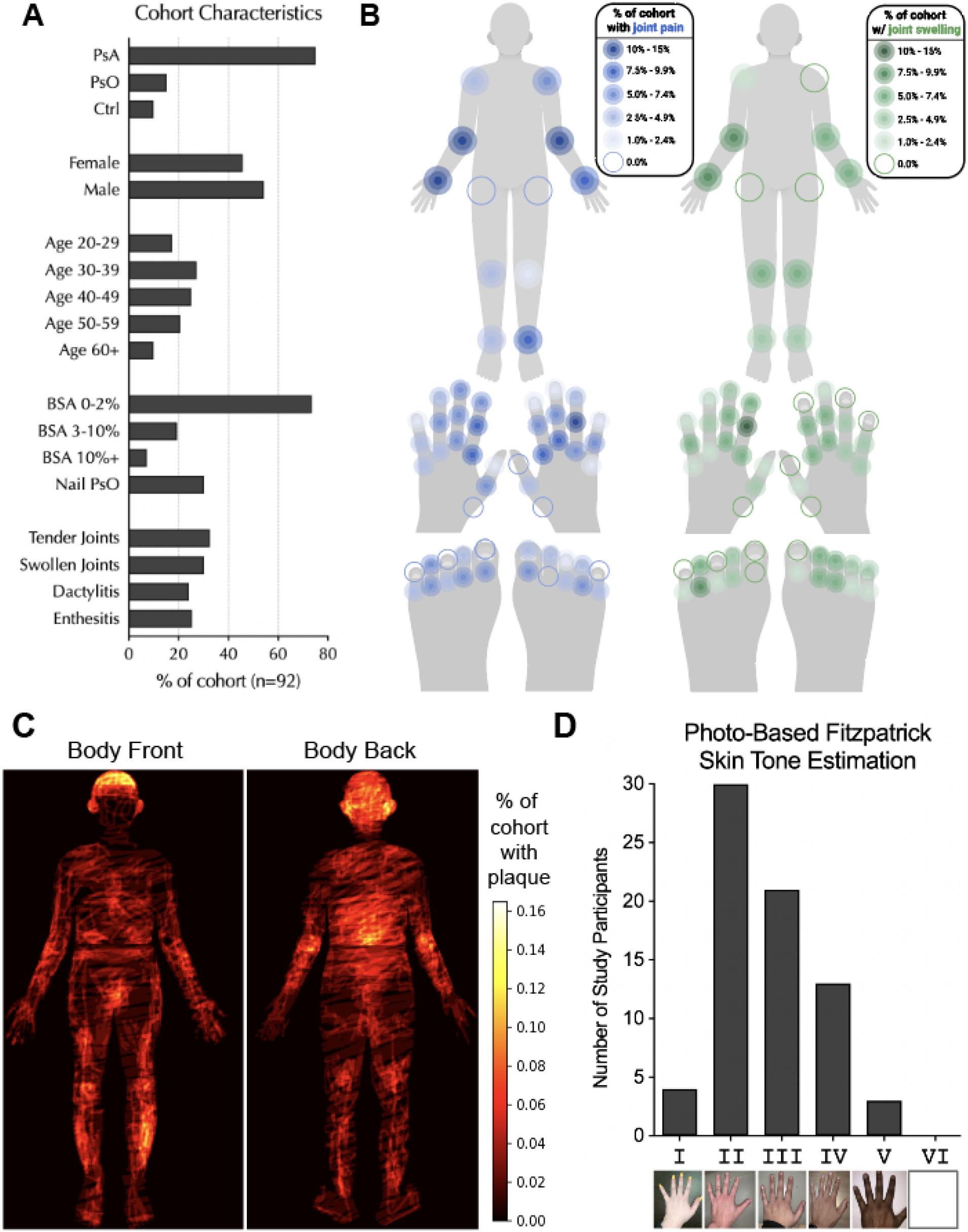
Cross-sectional validation study cohort characteristics. **(A)** Demographic and clinical characteristics of the validation study cohort (PsA = Psoriatic Arthritis, PsO = Psoriasis, Ctrl = Healthy Control, BSA = Body Surface Area). **(B)** Body map representation of clinically-assessed joint tenderness (left, blue) and joint swelling (right, green) percentage in overall cohort. **(C)** Body map representation self-identified plaque location percentage in overall cohort. **(D)** Cohort distribution of estimated Fitzpatrick skin tone based on novel method of hand image processing.

Clinical assessment of psoriasis typically includes an estimation of the percent body surface area (%BSA) that is affected by plaques and a Physician Global Assessment (PGA) consisting of a five-point rating scale of plaque characteristics along the axes of erythema, induration, and scaling.^45^ To investigate the clinical validity of the self-administered *Psoriasis Draw* measurement, a digital %BSA from drawn pixel calculations was compared to physician-assessed %BSA estimation performed during a concurrent physical exam (Fig. 2A). The digitally-acquired %BSA exhibited a very strong^46^ Lin’s concordance correlation coefficient (CCC) of 0·94 (0·91–0·96). We then assessed the extent to which a self-captured photo from the *Psoriasis Area Photo* assessment could recapitulate PGA ratings of the same plaque during a physical exam. Self-captured images were presented in a web-based portal and rated by a panel of physicians (n=5). Remote physician ratings were rarely off by more than 1 (Fig. 2C), demonstrating moderately strong^46^ concordance of CCC_erythema_=0·72 (0·59–0·85), CCC_induration_=0·72 (0·62–0·82), CCC_scaling_=0·60 (0·48–0·72). On average, the remote raters chose “can’t tell” for 11% of the images (Fig. 2D), which was relatively consistent across erythema, induration and scaling. These unratable images were primarily located on the scalp, suggesting that self-captured digital measures may not be feasible for all areas of the body.

**Fig. 2:**
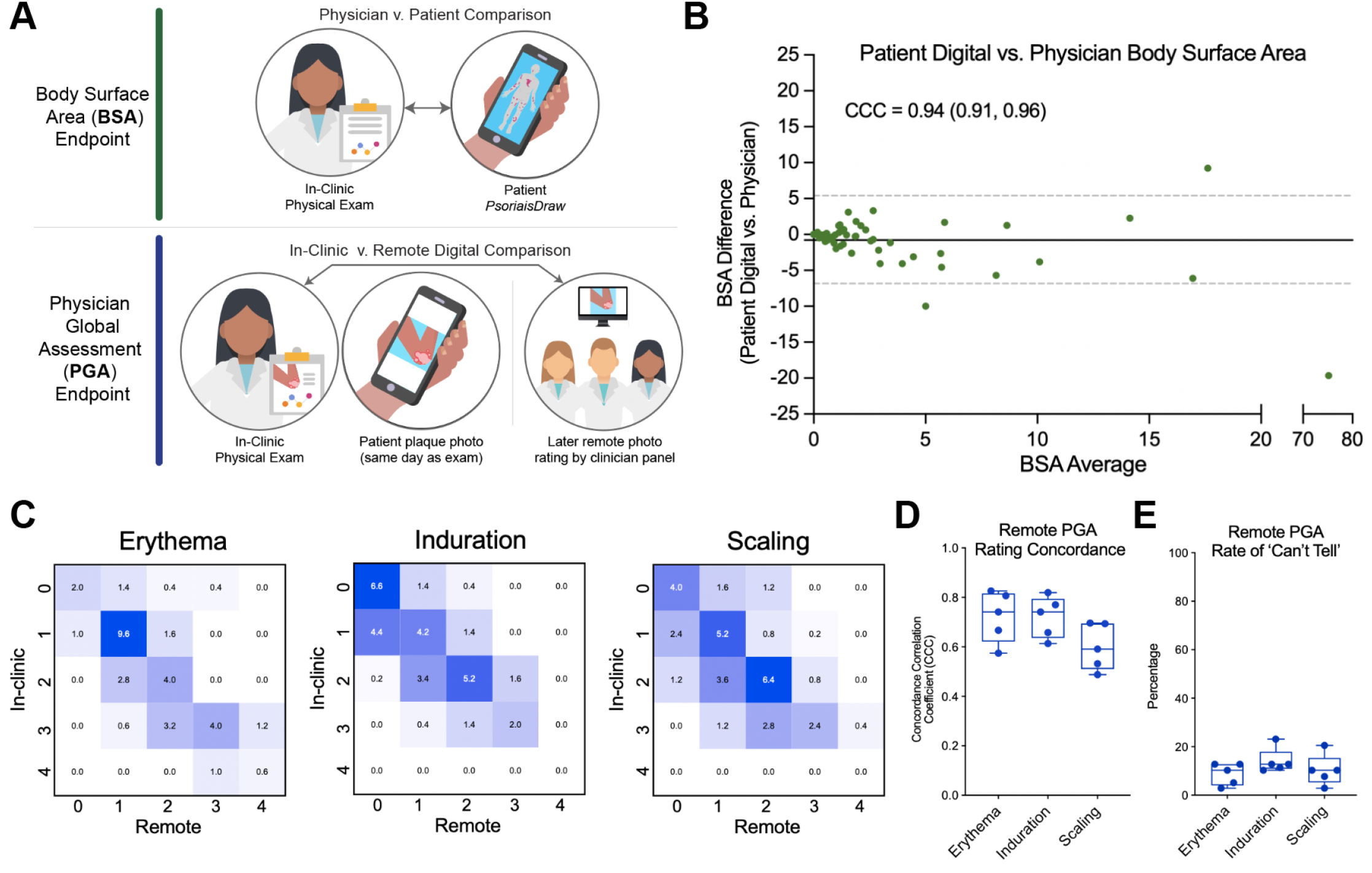
Clinical validation of digital assessments of cutaneous disease. **(A)** Validation study design for digital body surface area (BSA) and digital physician global assessment (PGA) endpoints **(B)** Bland-Altman plot comparing performance of patient digital BSA with physician in-clinic BSA (CCC = Lin’s Concordance Correlation Coefficient) **(C)** Confusion matrices of PGA scores for erythema, induration, and scaling between an in-clinic assessment and remote physicians. Decimal values in boxes are the average of 5 remote physician raters. **(D)** Concordance of each remote physician for each parameter of PGA **(E)** Rate of the annotation option ‘Can’t Tell’ for each remote physician rater.

Assessing musculoskeletal symptoms such as joint tenderness or enthesitis is essential for early detection of psoriatic arthritis. However, myalgias (i.e., muscle pain) and arthralgias (i.e., joint pain without inflammation) are common in the general population, leading to a difficulty for general practitioners^47^ and patients^48^ to correctly identify inflammatory-type musculoskeletal symptoms indicative of emerging PsA. To investigate the potential of self-administered digital measurements to detect psoriatic joint pain and enthesitis, we developed three assessments (*Painful Joints, Digital Jar Open, 30-Second Walk)* whose outputs were compared against in-clinic physical examination (Fig. 3A). Patient-reported joint pain from the *Painful Joints* digital assessment performed poorly (CCC = 0·33 (0·20–0·45)) compared to physician assessment of tender joints (Fig. 3B). These results are highly consistent with a previously reported comparison between patient and physician reported outcomes,^48^ confirming the difficulty of recapitulating physical exam findings for psoriatic joint tenderness with a patient self-reporting tool.

**Fig. 3:**
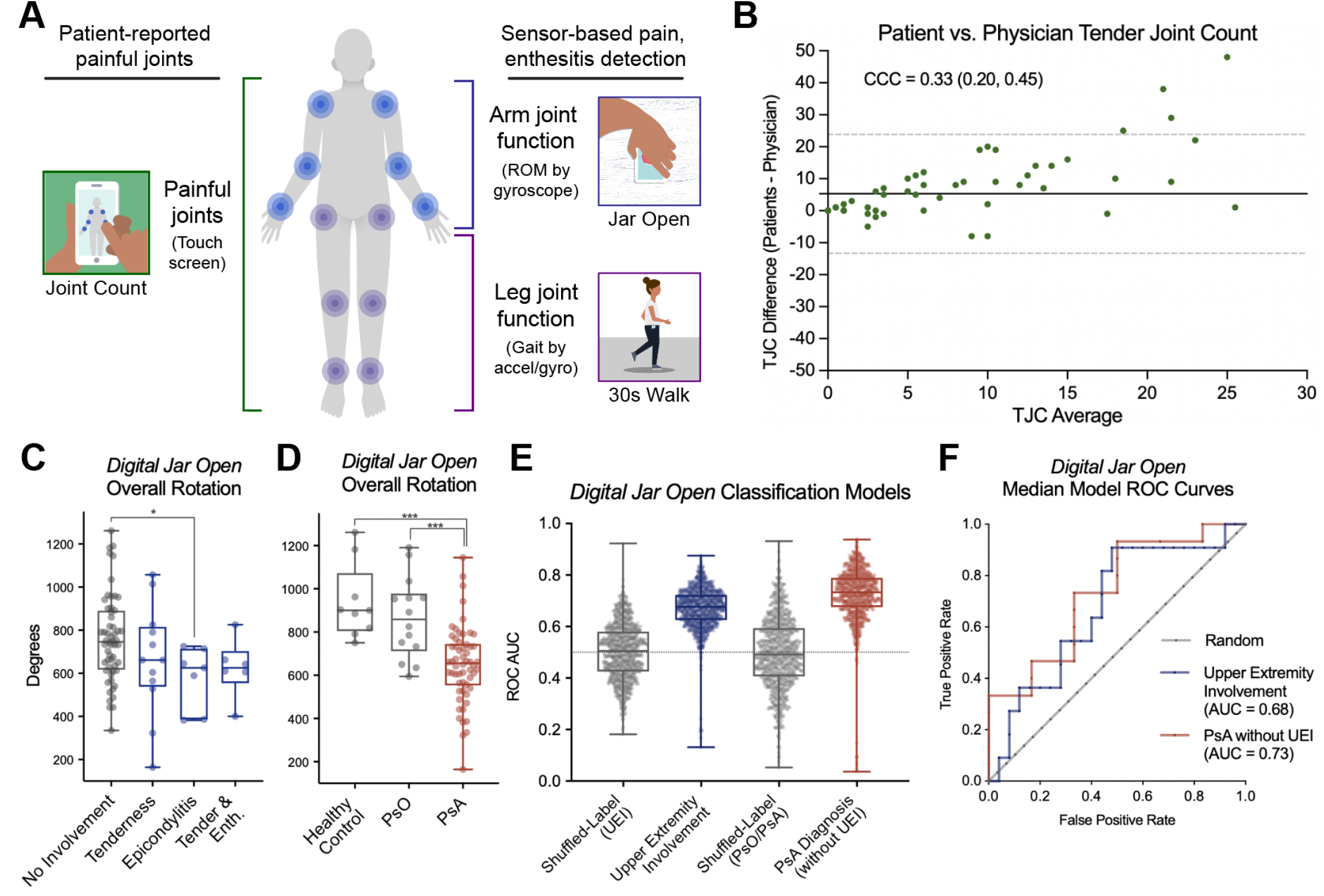
Sensor-based detection of joint tenderness, enthesitis, and psoriatic arthritis. **(A)** Graphical representation of *Psorcast* digital assessments to detect full-body joint pain (*Painful Joints)*, upper extremity involvement (*Digital Jar Open)* or lower extremity involvement (*30-Second Walk)* with joint tenderness or enthesitis **(B)** Bland-Altman plot comparing patient-reported painful joints with in-clinic tender joint count assessment. **(C)** Overall rotation endpoint from *Digital Jar Open* assessment in participant populations with no upper extremity involvement and those with upper extremity tender joints and/or enthesitis. **(D)** Overall rotation in healthy participants, and those with psoriasis (PsO) and psoriatic arthritis (PsA). **(E)** Performance of 1,000 iterations of classification model training using age-adjusted overall rotation feature. Labels are randomly shuffled to provide a matched control. (ROC AUC = Receiver-Operator Curve Area Under the Curve. **(F)** Median model performance (ROC) for detection of upper extremity involvement or identification of a psoriatic arthritis diagnosis in those without clinically diagnosed upper extremity involvement.

We hypothesized that sensor-based measurements of functional movement may detect psoriatic joint tenderness while also capturing functional impairment due to enthesitis. Expanding on a precedent of wrist pain quantification by smartphone range of motion measurement in rheumatoid arthritis,^25,26^ we sought to measure full-arm rotational motion to detect upper extremity involvement (UEI) that includes joint tenderness of the wrist, elbow, or shoulder as well as lateral epicondylitis.

Overall rotation from the *Digital Jar Open* measurement was sufficient as a single feature from the motion sensor data to distinguish populations with and without physician-assessed arm joint tenderness and/or enthesitis (Fig. 3C). Overall rotation was significantly decreased in those with psoriatic arthritis (PsA) compared with psoriasis or healthy controls (Fig. 3D). As we detected a confounding effect on overall rotation with age, we used age-adjusted overall rotation in our models to avoid overestimating predictive performance (appendix pp 9-26). Models for detecting UEI or identifying a PsA diagnosis in individuals without UEI performed markedly better than randomly shuffled labels (Fig. 3E). The median UEI classification model performed with an ROC AUC of 0·68 (0·67–0·68) (Fig. 3F), indicating that the self-administered *Digital Jar Open* assessment has the potential for recapitulating physical exam-based diagnosis of psoriatic joint tenderness of the wrist, elbow, or shoulder, or lateral epicondylitis. Interestingly, when we removed all healthy controls and individuals with clinically-assessed UEI from the cohort and analyzed whether the *Digital Jar Open* assessment could distinguish between psoriasis and psoriatic arthritis, we observed a similar model performance (median ROC AUC = 0·73). These results suggest that functional impairment of the upper extremities from psoriatic arthritis is detectable despite lack of concurrent clinical diagnosis of upper extremity joint tenderness or enthesitis.

We did not observe significant discriminatory capacity for lower extremity involvement from the *30-Second Walk* assessment features (appendix p 6). This may be due to the increased degrees of freedom during gait assessment from a phone in the pocket or waistband relative to a table-bound phone in the *Digital Jar Open* test, resulting in a much larger number of derived features and confounders. These results suggest that a larger cohort size will be needed to investigate the validity of this gait-based assessment for lower extremity involvement.

Nail psoriasis is a common clinical phenotype of psoriasis that may serve as a predictor of progression to psoriatic arthritis.^49^ To investigate the potential for automated detection of nail psoriasis using patient-provided hand image data, we developed an image processing workflow including a nail object detection model and deep learning-based image classification model of nail psoriasis. Model predictions were then evaluated relative to in-person physical exam results and compared to remote evaluation of nail images by a panel of physicians (Fig. 4A).

**Fig. 4:**
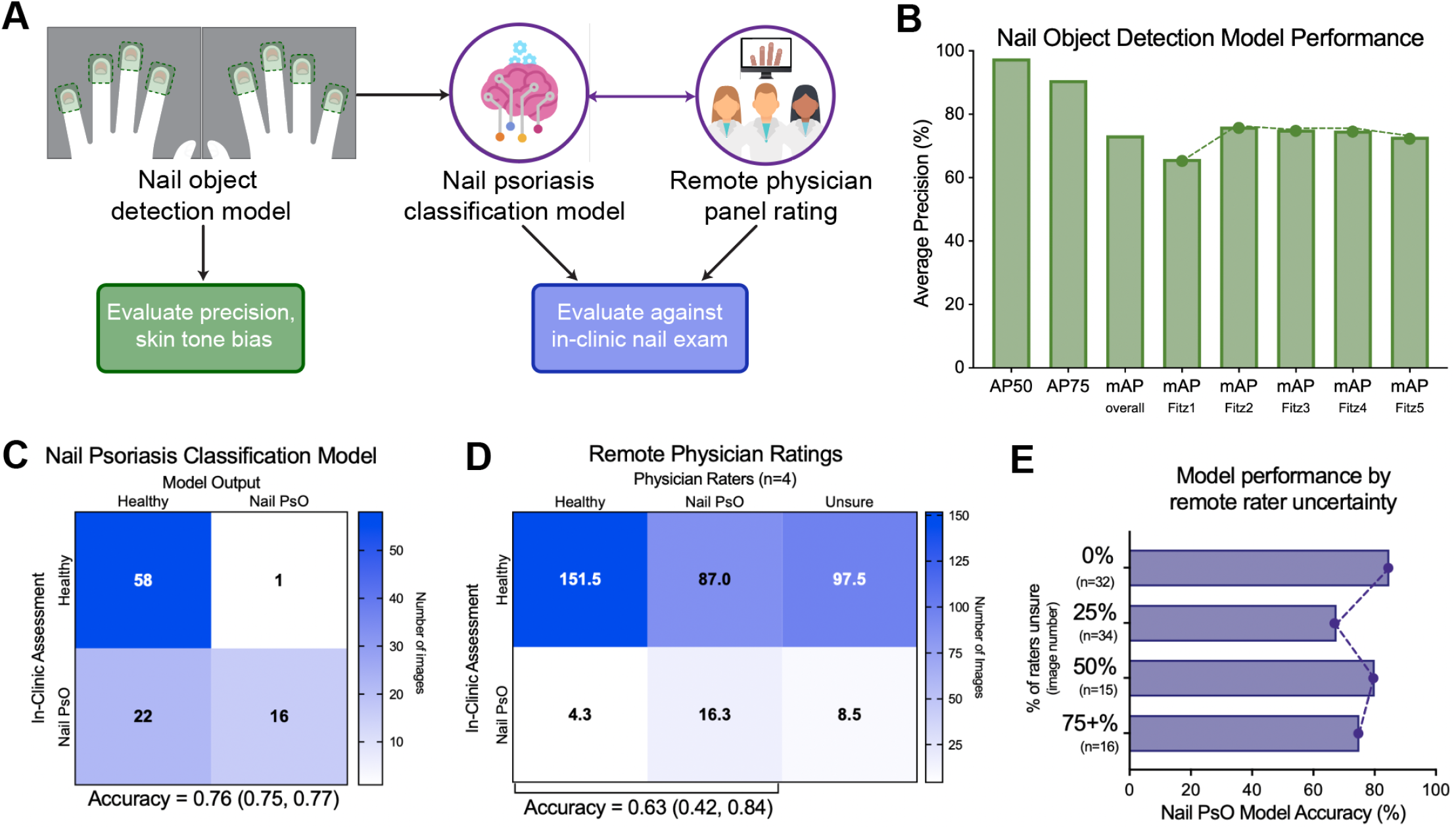
Detection of nail psoriasis from patient-captured hand images. **(A)** Image processing and machine learning evaluation workflow for patient hand imaging. **(B)** Precision of nail object detection model overall and within each estimated Fitzpatrick skin tone. (AP50 = Average precision for intersection over union (IoU) of 0.5; AP75 = average precision for IoU=0.75, mAP = mean average precision) **(C)** Confusion matrix for nail psoriasis model relative to in-clinic assessment. **(D)** Confusion matrix for remote physician rating of nail psoriasis relative to in-clinic assessment. **(E)** Performance of nail psoriasis model in cases where varying fractions of the remote physician panel were unsure of the presence or absence of nail psoriasis.

Nail object detection demonstrated a mean average precision (mAP) of 73·4%, with highly similar performance across estimated Fitzpatrick skin tones (Fig. 4B). The nail psoriasis classification model exhibited an accuracy of 0·76 when evaluating nail images with ground-truth labels from in-clinic physical exams (Fig. 4C). These results provide preliminary evidence that patient-provided hand images contain sufficient information to detect nail psoriasis using deep learning models.

A remote panel composed of 4 specialized physicians (i.e., dermatologists) rating the full set of nail images from the in-clinic cohort demonstrated an average accuracy of 0·63. However, this accuracy did not reflect the additional options for physicians to classify images with ‘unsure’ or ‘not of sufficient quality’, which occurred on average in 29% of cases (Fig. 4D). When comparing the nail classification model with remote physician raters, we observed that physicians had a higher average recall of nail psoriasis (0·56) than the model (0.42), while the model had a higher precision (0·93) than physician raters (0·16) (Table 1). We next sought to explore the performance of the nail classification model in cases where physicians expressed uncertainty. Model accuracy remained consistent in spite of an increasing fraction of physicians rating ‘unsure’ (Fig. 4E), which is a similar finding from recent efforts to employ deep learning-based models for differential diagnosis of skin disease.^50^

## Discussion

Here we describe the evaluation of a set of novel, smartphone-based assessments for patient self-measurement of psoriatic disease phenotypes compared to clinical assessments by a specialized physician. These measures for cutaneous plaques, peripheral arthritis, and nail psoriasis were validated in a cross-sectional setting utilizing rheumatology and dermatology experts in psoriatic disease. Taken together, these results demonstrate the validity of digital measurements as a new method for remote, self-assessed psoriatic disease activity monitoring.

While applications have been developed for digitally tracking psoriatic disease,^51–53^ they largely focus on questionnaire-based patient reported outcomes. To our knowledge, ours is the first application using ubiquitous smartphone sensor technology for patients to remotely measure their psoriatic disease phenotypes, including detection of nail psoriasis and a continuous variable outcome measure of joint tenderness and enthesitis based on range of motion (ROM). While this ROM-based *Digital Jar Open* assessment was only evaluated in a cross-sectional setting for a single time point, it has the potential to remotely capture granular changes of functional impairment over time. Interestingly, we observed that the *Digital Jar Open* assessment, while only measuring rotation of the upper extremities, was able to classify an overall PsA diagnosis despite no concurrent clinically-assessed upper extremity involvement with tender joints or enthesitis. While this model classification is not as useful in patients with known, pre-existing damage that has already impaired range of motion, it could be valuable for those at high risk for PsA as a scalable screening tool to guide the use of clinical imaging modalities for early PsA detection.^54^

The product of physician’s global assessment and body surface area (PGA×BSA) is increasingly used as a simpler and highly correlated alternative to the widely used PASI score for assessment of psoriasis.^55,56^ The *Psorcast* app utilizes two digital measurements, *Psoriasis Area Photo* and *Psoriasis Area Draw*, that are designed to measure analogous endpoints to PGA and BSA, respectively. The innovation of these digital assessments is the ability for patients to remotely and rapidly self-measure their psoriasis with commonly used smartphone interactions (a ‘selfie’ and a drawing). The PGA component of this digital PGAxBSA approach currently relies on remote assessments by specialists to determine plaque characteristics. This has been shown to be an effective approach for care,^57^ however, the PGA may be a task that can be augmented with machine learning in the future using larger validation datasets.^58^

These results should be interpreted in light of their limitations. The present study had a limited sample size, which is an important caveat to the results from our motion sensor and image-based machine learning models. These should be considered encouraging, proof-of-concept findings that provide the rationale for much larger studies to assess robustness and generalizability of the models to a larger population. Larger sample sizes may also be required to more definitely assess the potential of gait dynamics to detect lower extremity involvement with tender joints and enthesitis or image-based assessment of tender joints or dactylitis.

In this study, we focused on patients being seen by psoriatic disease experts in academic centers, which may capture a distinct population of patients that may not necessarily be generalizable. For example, our cohort largely lacks individuals presenting with psoriatic plaques at the highest severity on the PGA rating scale, and individuals with the lightest and darkest skin tones. Despite the expertise of physicians conducting this study, it can be difficult to fully account for possible clinical confounders like superimposed, non-inflammatory pain syndromes (e.g., fibromyalgia, osteoarthritis, and mechanical injury). Lastly, we describe physician-conducted physical examinations and remote image assessment as comparators, but we recognize that in-clinic examination was restricted to the captured plaque and did not represent a full body evaluation, that physicians and patients often disagree about the state of their disease,^59^ and that there is no validated, objective gold standard for many of these assessments.

The use and continued development of this technology opens up new possibilities for both clinical care and research endeavors on a large scale. Clinically, the recent COVID-19 pandemic has highlighted even further challenges related to access to care and the role that technology can play to fill that gap. While this application was not conceived as a potential replacement for physician visits, digital biomarkers offer novel avenues for patients to have an ever-increasing role in their care, while tracking their skin and joint symptoms for prospective fluctuation in disease activity. Additionally, as psoriatic disease can be episodic and flares are difficult to track, patients could more easily present data to their physicians about symptoms and disease activity that occurred in-between visits, and modulate the frequency of appointments accordingly.^60^ *Psorcast* measurements are currently being validated for their ability to assess disease changes longitudinally, allowing for more frequent symptom monitoring in clinical trials, more granular insight into the time course of medication action, and possible integration into precision medicine strategies aimed at identifying responders from non-responders to specific therapies.

Here, we provide the groundwork for patient-driven, remote measurement of psoriatic disease. Importantly, to facilitate external validation of these results while promoting wide adoption of these instruments, smartphone software and analysis pipelines for the *Psorcast* suite have been made freely available to the scientific community (see *Data Sharing* section). The *Psorcast* measurements, combined with passive and contextual measurements of environmental variables and lifestyle factors such as diet, physical activity, and sleep will form the basis for a large-scale, longitudinal study with an increased sample size and more diverse patient population to refine and further validate benchmark digital measurements and model performance. Ultimately, we seek to create an integrated measurement tool to allow timely disease monitoring in real-world settings and an analytical framework for ‘psoriatic disease forecasts’ for prediction of therapeutic response, flare/remission cycles, and early detection of the transition from psoriasis to psoriatic arthritis.

## Data Availability

De-identified validation study datasets that can be shared publicly are available on the Synapse platform at the url: https://www.synapse.org/#!Synapse:syn26840742/wiki/ Guidelines to reproduce the analysis results and figures is shared in this Github Repository: https://github.com/Sage-Bionetworks/psorcast-validation-manuscript. App software can be found at https://github.com/Sage-Bionetworks/Psorcast-iOS.

https://github.com/Sage-Bionetworks/psorcast-validation-manuscript

https://github.com/Sage-Bionetworks/Psorcast-iOS

## Contributors

DEW, RHH, LMPC, WM, MD, SMR, AO, MRK, LMM, SKS, LO, JFM and JUS conceptualized and designed the study. RHH, LMPC, SC, AN, CC, JFM and JUS enrolled and examined study participants. DEW, RHH, LMPC, WM, MD, ES, JFM, and JUS developed the smartphone application and measures. DEW, MT, AT, VY, ECN performed data analysis and model development. RHH, LMPC, WF, MN, MY, RLC, AN, DY, AK and JFM performed remote image annotation. DEW, RHH, LMPC, MT, AT, VY, ECN, SMR, AO, AK, LMM, SKS, LO, JFM, JUS drafted the manuscript and performed critical revision. All authors contributed to data interpretation and read and approved the final manuscript.

## Acknowledgements

This work was supported by contributions from the pre-competitive Psorcast Digital Biomarker Consortium partner institutions. Within the consortium, we would particularly like to acknowledge the efforts of Bhumik Parikh, ‘Matladi N. Ndlovu, Pamela Young, Soumya Chakravarty, and Amanda Pacia. We would also like to acknowledge the supporting efforts of Stockard Simon, Phil Snyder, Vanessa Barone, Erin Mounts, Shannon Young, and Alx Dark.

