## Supplementary Appendix for "Clinical validation of digital biomarkers and machine learning models for remote measurement of psoriasis and psoriatic arthritis"

##### Table of Contents

**Supplementary Figures**

Supplementary Figure 1. *Psoriasis Draw* user interface 2
Supplementary Figure 2. *Psoriasis Area Photo* user interface 2
Supplementary Figure 3. *Digital Jar Open* user interface 3
Supplementary Figure 4. *Fingers Photo* user interface 3
Supplementary Figure 5. *Toes Photo* user interface 4
Supplementary Figure 6. *30-Second Walk* user interface 4
Supplementary Figure 7. *Painful Joint Count* user interface 4
Supplementary Figure 8. Patient/physician impact matrix for assessments 5
Supplementary Figure 9. *30-Second Walk* exploratory analysis 6

**Supplementary Tables**

Supplementary Table 1. Validation study cohort demographics and diagnoses 7-8

**Supplementary Methods**

*Digital Jar Open* confounding variable analysis 9-26
Nail object detection and nail psoriasis classification 27-32
Hand image processing and Fitzpatrick skin tone estimation 33-37

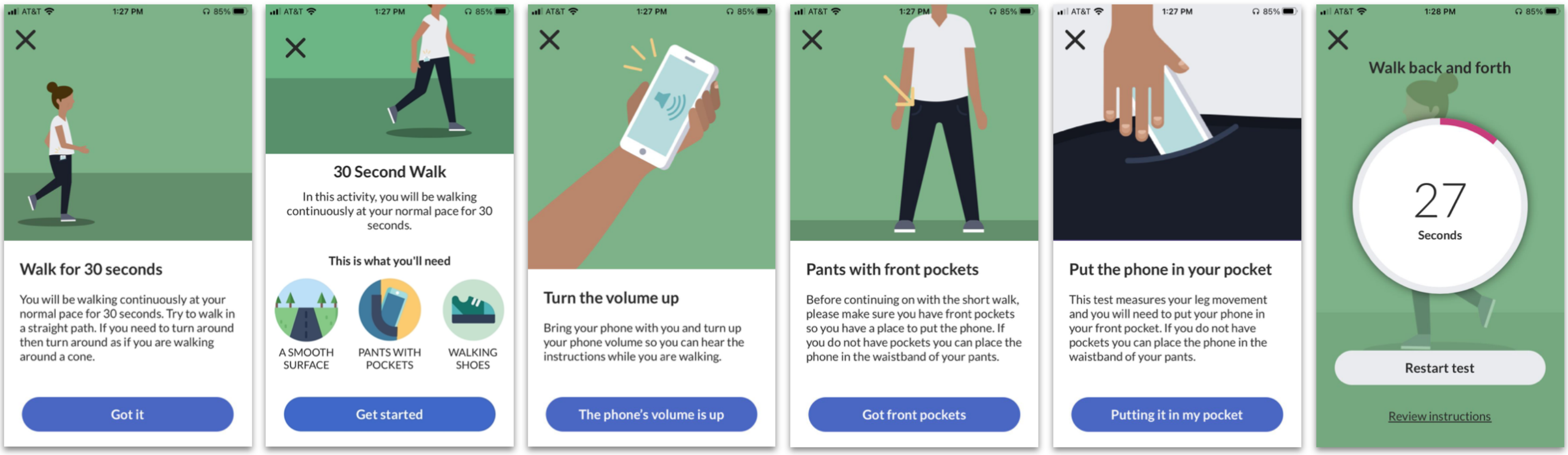


**Supplementary Figure 1.** *Psoriasis Draw* **user interface.** Participants are first provided with an option to select the overall areas of the body that are affected by psoriasis. They are then provided with an outline of a human figure for each region selected and are instructed to draw in the area and extent of their active skin psoriasis. This then calculates an estimated percent body surface area based on the percentage of pixels 'drawn' out of the total amount of pixels within the body map.

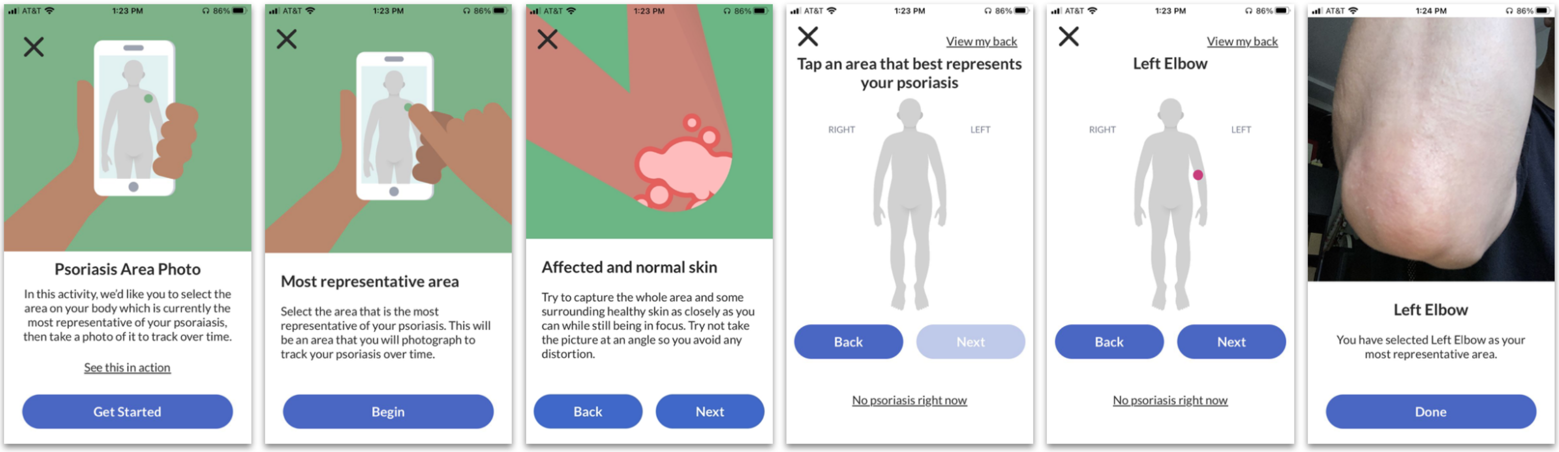


**Supplementary Figure 2.** *Psoriasis Area Photo* **user interface.** This activity accesses the smartphone camera to allow for capture of skin disease. Participants are instructed to pick an area of their skin, affected by psoriasis, that best represents their skin disease or is mostly commonly involved. They are provided with a human outline and via touch-screen mark off the area of the body that they will be capturing with the photo. They are then instructed to take a picture of the area and have the ability to retake the photo as many times as needed to capture the area adequately.

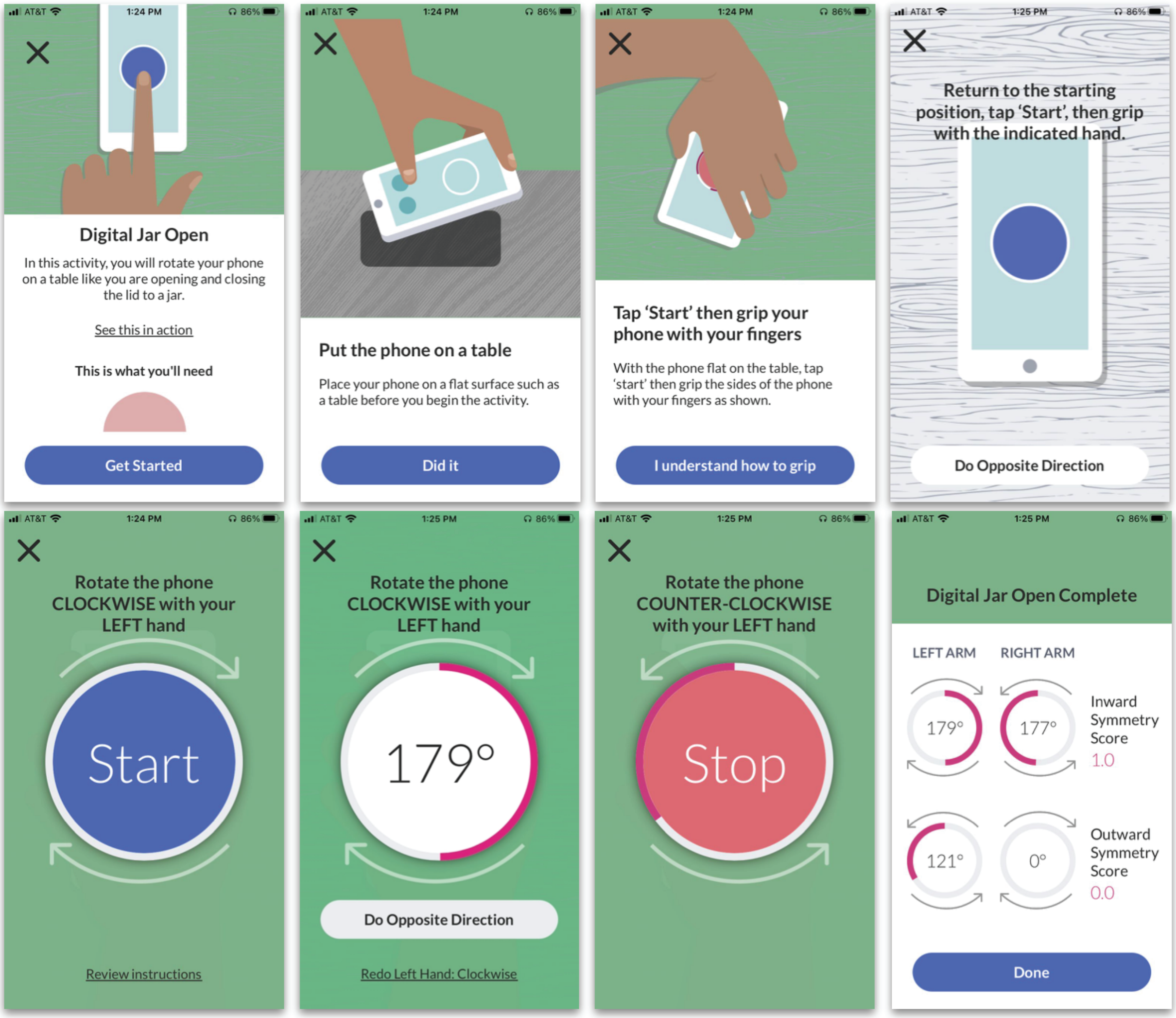


**Supplementary Figure 3.** ***Digital Jar Open*** **user interface.** This assessment employs smartphone motion sensors (accelerometers and gyroscope) to measure the range of motion of the upper extremities during inward and outward rotation. Participants are instructed to grasp the smartphone in one hand and rotate the phone as far as possible both internally and externally while the phone is placed on a flat table. The rotational movement engages the shoulder, elbow, and wrists. Degrees of rotation are recorded in each direction and this process is repeated using both the left and right sides.

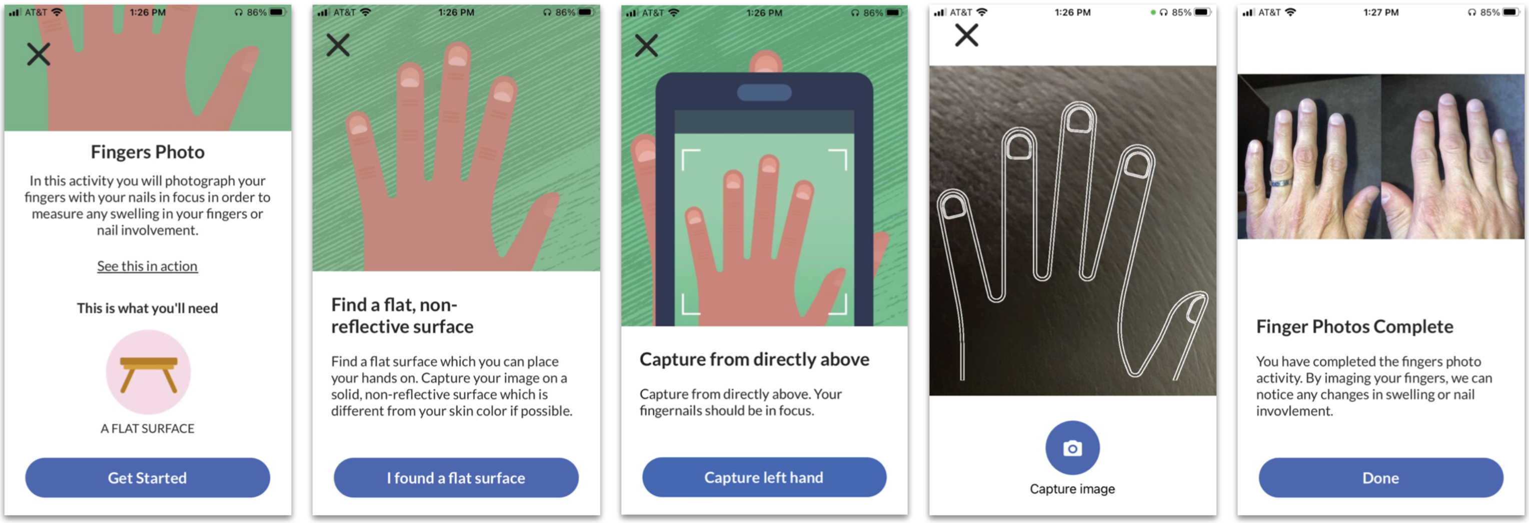


**Supplementary Figure 4.** ***Fingers Photo*** **user interface.** This assessment accesses the smartphone camera. Patients are asked to align their hand within an outline form and assure that their nails are in focus in the captured image. The image capture is then repeated for the other hand.

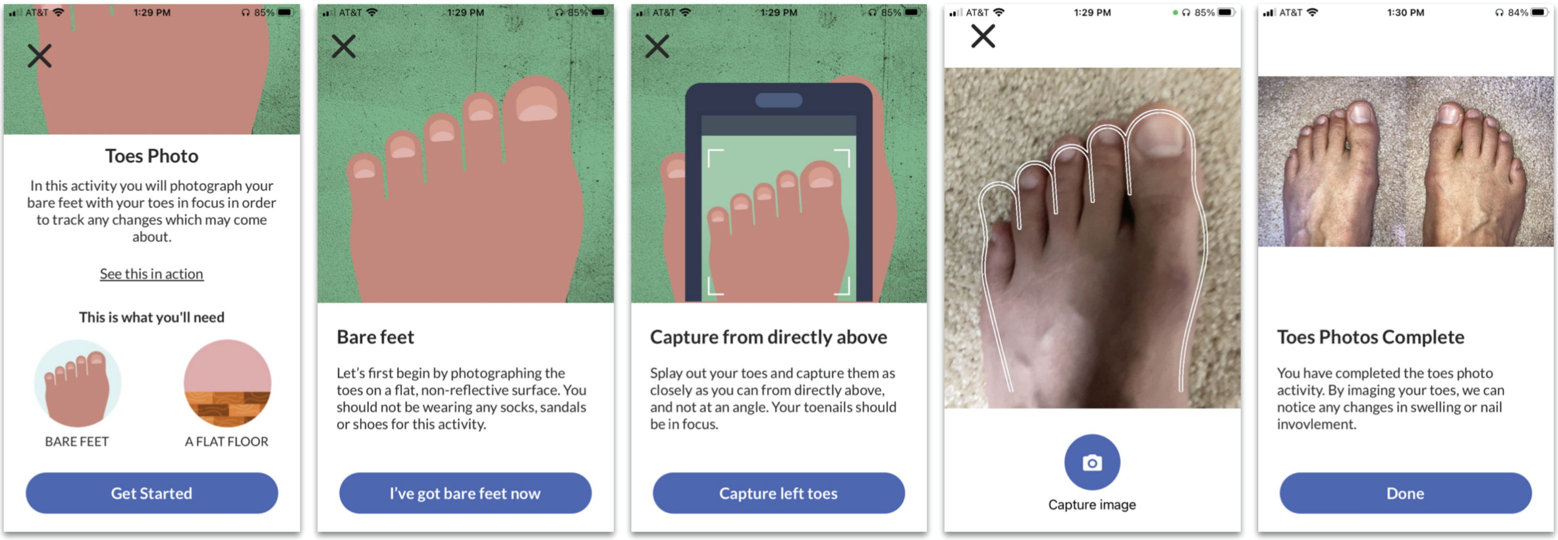


**Supplementary Figure 5. *Toes Photo* user interface.** This assessment accesses the smartphone camera. Patients are asked to remove their shoes and socks and align a foot within an outline form and assure that their nails are in focus in the captured image. The image capture is then repeated for the other foot.

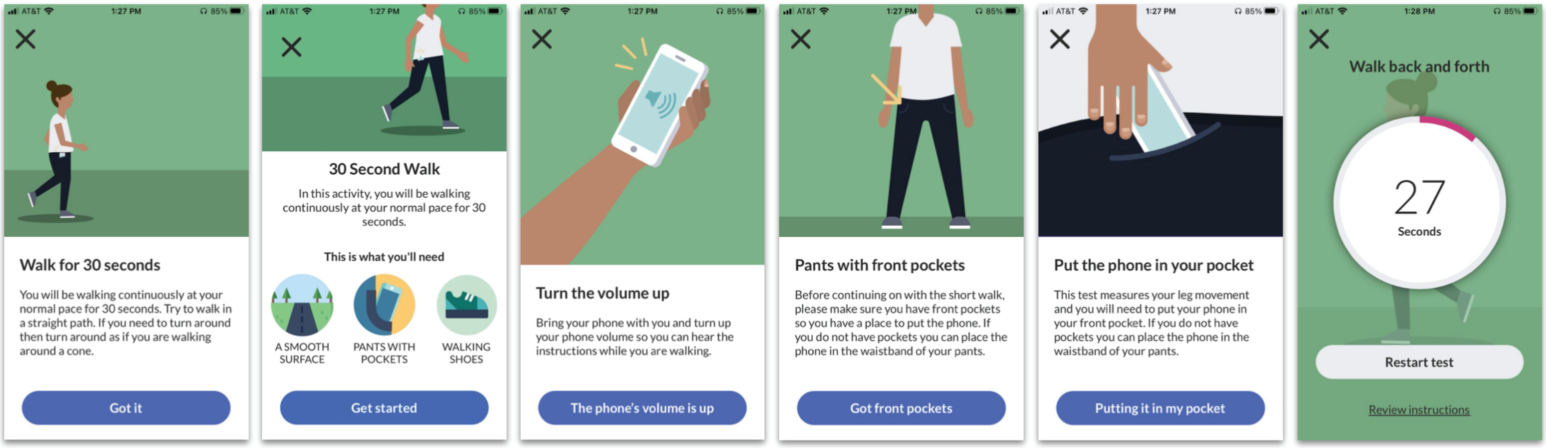


**Supplementary Figure 6. *30-Second Walk* user interface.** This assessment engages the smartphone gyroscope/accelerometer to measure gait dynamics. Patients are asked to place the phone in their front pocket and walk for 30-seconds at their normal pace.

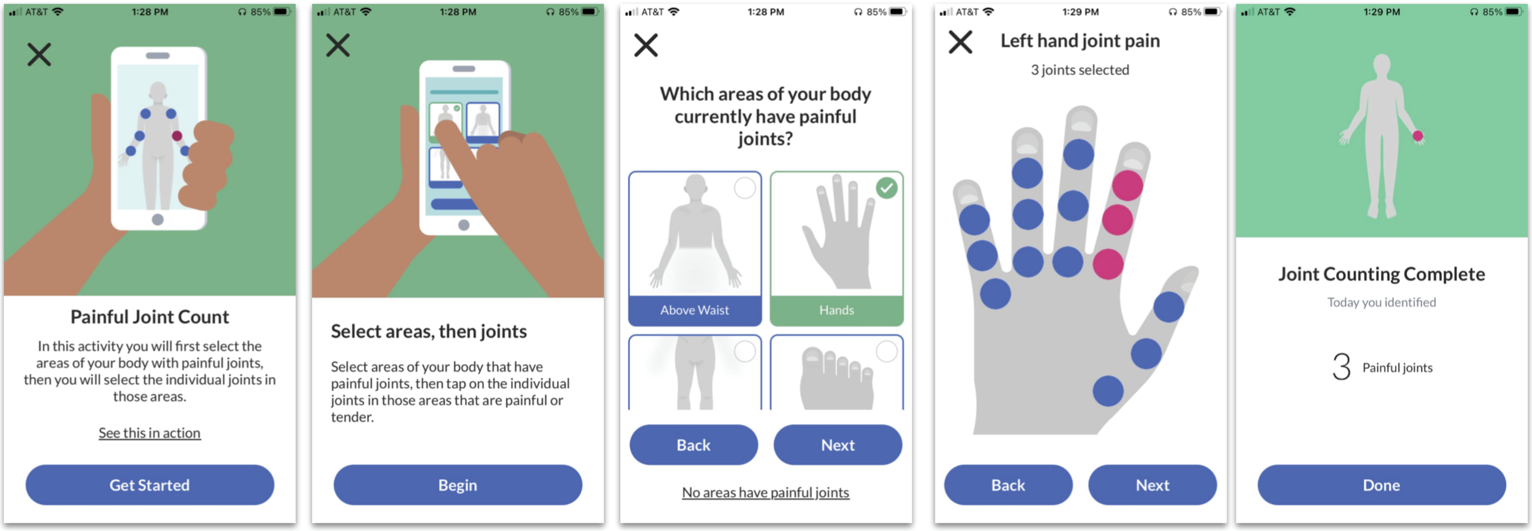


**Supplementary Figure 7. *30-Second Walk* user interface.** Participants are first provided with an option to select the overall areas of the body that are affected by painful joints. They are then provided with an outline of a human figure for each region selected and are instructed to select from the pre-specified joints that are causing them pain and/or discomfort.

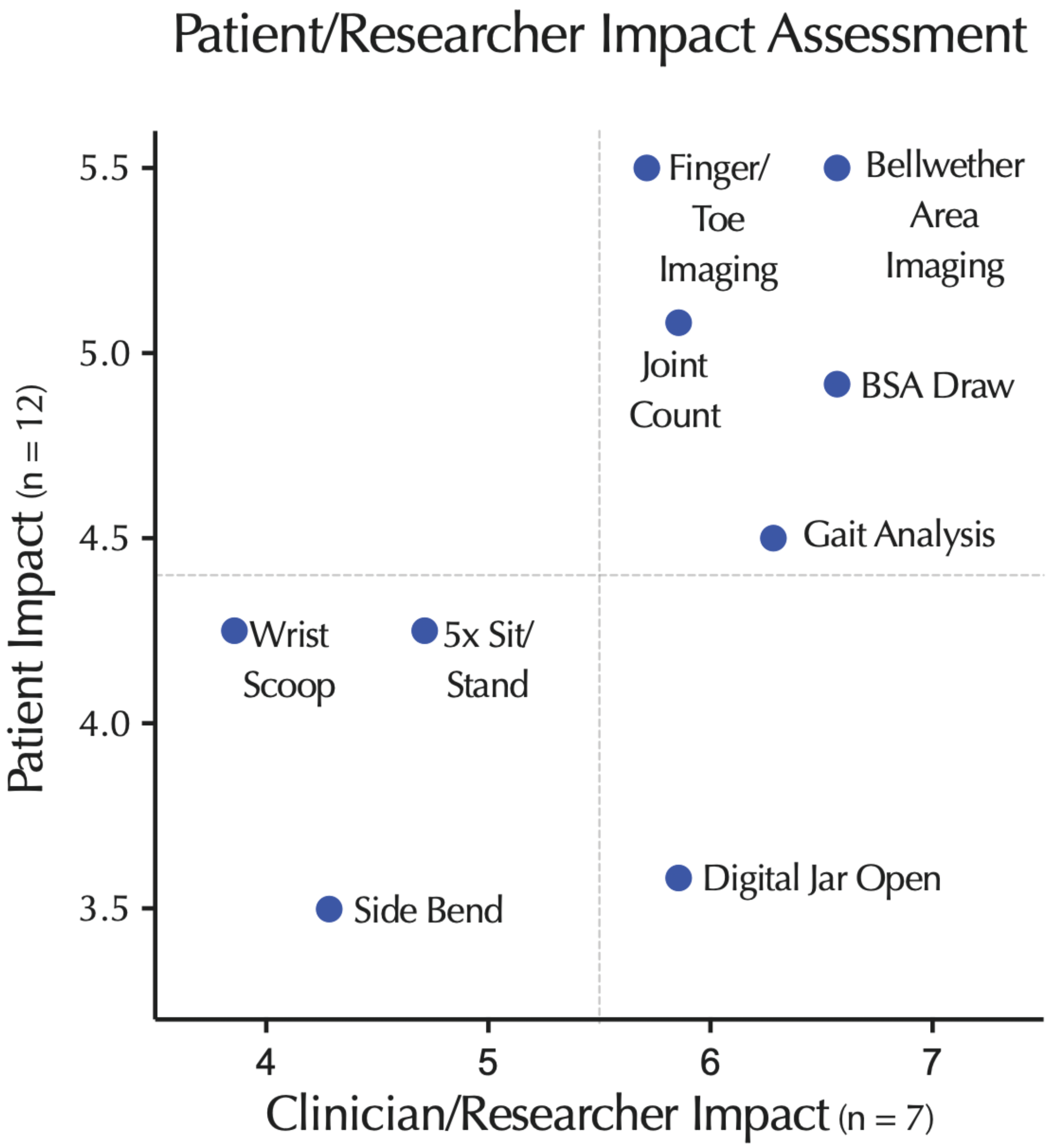


**Supplementary Figure 8. Patient/physician impact matrix for proposed assessments.** Foundational user reserach employing ranked choice voting survey responding to the prompt, “Please rank the following according to the impact they would have for you as a [patient/physician]”. The choices in this survey were accompanied by a user interface illustrating the function of the measure.

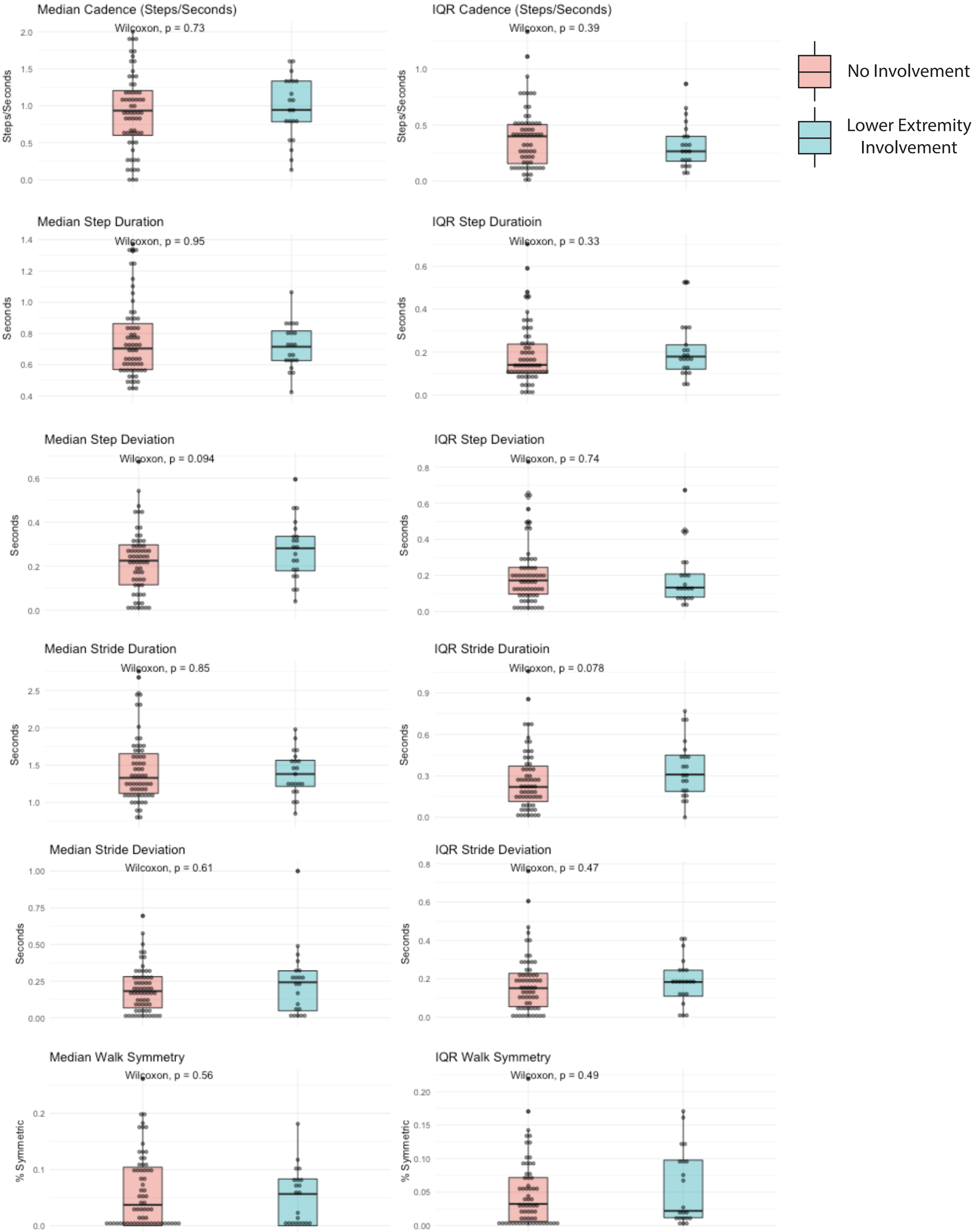


**Supplementary Figure 9. 30-Second Walk exploratory analysis.** Exploratory analysis of *30-Second Walk* assessment data for separation between participants with and without lower extremity involvement with tender joints or enthesitis.

| Participant Identifier | Diagnosis | Visit | Age Decile | Sex | BSA (%) | Nail PsO (Fingers) | Nail PsO (Toes) | Tender Joints | Swollen Joints | Enthesitis Label | Dactylitis (Fingers) | Dactylitis (Toes) |
|---|---|---|---|---|---|---|---|---|---|---|---|---|
| SITE1_001 | Control | 1 | 30s | Male | 0 | None | None | 0 | 0 | None | None | None |
| SITE1_002 | Control | 1 | 30s | Female | 0 | None | None | 0 | 0 | None | None | None |
| SITE1_003 | PsO | 1 | 20s | Female | 4 | None | None | 0 | 0 | None | None | None |
| SITE1_004 | PsA | 1 | 40s | Male | 3 | L4 | None | 0 | 4 | left_achilles | None | R3 |
| SITE1_005 | Control | 1 | 40s | Male | 0 | None | None | 0 | 0 | None | None | None |
| SITE1_006 | Control | 1 | 30s | Female | 0 | None | None | 0 | 0 | None | None | None |
| SITE1_007 | Control | 1 | 40s | Female | 0 | None | None | 0 | 0 | None | None | None |
| SITE1_008 | PsA | 1 | 20s | Male | 1 | None | None | 0 | 0 | None | None | None |
| SITE1_009 | Control | 1 | 20s | Female | 0 | None | None | 0 | 0 | None | None | None |
| SITE1_010 | PsA | 1 | 40s | Male | 1 | None | None | 10 | 0 | None | None | None |
| SITE1_011 | PsO | 1 | 40s | Female | 0 | None | None | 0 | 0 | None | None | None |
| SITE1_012 | PsA | 1 | 20s | Female | 3 | L1, L2, L3 | None | 4 | 6 | None | none | L4, R4 |
| SITE1_013 | PsA | 1 | 40s | Male | 1 | None | L1, R1 | 1 | 2 | None | None | None |
| SITE1_014 | PsA | 1 | 40s | Female | 0.5 | L3 | None | 2 | 2 | None | None | None |
| SITE1_015 | PsO | 1 | 20s | Male | 0.5 | None | None | 0 | 0 | None | None | None |
| SITE1_017 | PsA | 1 | 30s | Male | 1 | L1, L2, L5, R4 | None | 0 | 0 | None | None | None |
| SITE1_018 | PsA | 1 | 50s | Female | 1 | None | None | 8 | 8 | left_arm, right_arm | None | L2 |
| SITE1_019 | PsA | 1 | 30s | Male | 0 | None | None | 1 | 0 | None | None | None |
| SITE1_020 | PsA | 1 | 50s | Male | 0.5 | None | None | 0 | 0 | None | None | L3 |
| SITE1_021 | PsA | 1 | 30s | Female | 8 | None | None | 0 | 0 | right_achilles | L3, L4, L5 | L2 |
| SITE1_022 | PsA | 1 | 50s | Male | 0 | None | None | 0 | 0 | None | None | None |
| SITE1_023 | PsA | 1 | 30s | Male | 0 | None | None | 0 | 0 | None | None | None |
| SITE1_025 | PsA | 1 | 20s | Male | 2 | None | None | 0 | 0 | None | None | L5 |
| SITE1_027 | PsA | 1 | 50s | Female | 1.5 | None | None | 0 | 0 | None | None | None |
| SITE1_028 | PsA | 1 | 50s | Female | 0 | None | Not assessed | 0 | 0 | right_arm | None | None |
| SITE1_029 | PsO | 1 | 20s | Female | 1 | Not assessed | Not assessed | 4 | 4 | None | None | None |
| SITE1_030 | PsA | 1 | 20s | Female | 2 | None | None | 0 | 0 | None | None | R3, R4 |
| SITE1_031 | PsA | 1 | 50s | Female | 0 | None | L1, R1 | 0 | 0 | left_arm, right_arm | None | None |
| SITE1_032 | PsA | 1 | 30s | Male | 0 | L1, R1 | None | 0 | 0 | None | L1 | L2, R4, R5 |
| SITE1_033 | PsA | 1 | 30s | Male | 20 | Not assessed | None | 0 | 0 | left_arm, right_arm, left_achilles, right_achilles | None | None |
| SITE1_034 | PsA | 1 | 50s | Female | 0 | None | None | 0 | 0 | None | None | None |
| SITE1_035 | PsO | 1 | 20s | Male | 1 | None | None | 0 | 0 | None | None | None |
| SITE1_036 | PsA | 1 | 40s | Male | 0.5 | Not assessed | None | 0 | 0 | None | None | None |
| SITE1_037 | PsA | 1 | 50s | Female | 2 | None | L1, L2, L3, L4, L5, R1, R2, R3, R4, R5 | 0 | 0 | None | None | None |
| SITE1_038 | PsA | 1 | 60s | Male | 3 | None | None | 0 | 0 | None | L3 | None |
| SITE1_039 | PsA | 1 | 20s | Male | 0 | None | L1, R1 | 0 | 8 | left_achilles, right_achilles | None | L4, R1, R3, R4 |
| SITE1_040 | Control | 1 | 20s | Female | 0 | None | None | 0 | 0 | None | None | None |
| SITE1_041 | PsA | 1 | 40s | Male | 0 | L1, L2, L3, L4, L5, R1, R2, R3, R4, R5 | L1, L2, L3, L4, L5, R1, R2, R3, R4, R5 | 1 | 1 | None | None | none |
| SITE1_042 | PsA | 1 | 30s | Male | 0.5 | L1, L2, L3, L4, L5, R1, R2, R3, R4, R5 | L1 | 18 | 18 | right_achilles | None | L3, L4 |
| SITE1_043 | PsA | 1 | 40s | Male | 0 | None | None | 0 | 0 | None | None | None |
| SITE1_044 | PsA | 1 | 40s | Male | 0.5 | None | None | 0 | 0 | None | None | None |
| SITE1_045 | PsA | 1 | 60s | Female | 1.5 | None | None | 0 | 0 | right_arm, left_arm, right_knee, left_knee | None | None |
| SITE2_001 | PsO | 1 | 30s | Female | 1 | None | None | 0 | 2 | None | None | None |
| SITE2_002 | PsA | 1 | 40s | Male | 1 | None | None | 5 | 0 | None | None | None |
| SITE2_003 | PsA | 1 | 30s | Male | 0 | None | None | 1 | 5 | right_arm | None | None |
| SITE2_004 | PsA | 1 | 60s | Male | 0.5 | L1, L2, L3, L4, L5, R1, R2, R3, R4, R5 | L1, L2 | 13 | 9 | left_arm, right_arm | None | None |
| SITE2_005 | PsA | 1 | 50s | Male | 0 | None | None | 6 | 3 | None | None | None |
| SITE2_006 | PsO | 1 | 20s | Female | 13 | None | None | 0 | 0 | None | None | None |
| SITE2_007 | PsA | 1 | 20s | Female | 3 | None | None | 0 | 0 | None | None | None |
| SITE2_008 | PsA | 1 | 70s | Female | 0 | None | None | 13 | 9 | left_arm, right_knee | None | None |
| SITE2_009 | PsA | 1 | 40s | Female | 12 | None | None | 4 | 4 | right_achilles | None | None |

| | | | | | | | | | | | | |
|---|---|---|---|---|---|---|---|---|---|---|---|---|
| SITE2_010 | PsA | 1 | 40s | Female | 85 | None | None | 10 | 8 | left_arm, right_arm, right_knee | None | None |
| SITE2_012 | PsO | 1 | 60s | Male | 2 | None | None | 0 | 0 | None | None | None |
| SITE2_013 | PsA | 1 | 30s | Female | 4 | R4, L3 | None | 6 | 0 | left_arm | None | None |
| SITE2_014 | PsA | 1 | 40s | Female | 1 | None | L1 | 6 | 0 | None | None | None |
| SITE2_015 | PsO | 1 | 60s | Female | 8 | None | None | 0 | 0 | None | None | None |
| SITE2_016 | Control | 1 | 40s | Male | 0 | None | None | 1 | 1 | None | None | None |
| SITE2_017 | PsA | 1 | 50s | Female | 5 | None | None | 4 | 5 | None | None | None |
| SITE2_018 | Control | 1 | 30s | Female | 0 | None | None | 0 | 0 | None | None | None |
| SITE2_019 | PsA | 1 | 60s | Female | 1 | None | None | 0 | 0 | None | None | None |
| SITE2_020 | PsA | 1 | 30s | Female | 2 | None | None | 25 | 14 | left_arm, right_arm, left_knee, right_knee, left_achilles, right_achilles | None | None |
| SITE2_021 | PsO | 1 | 50s | Female | 1 | L1, L3, L4, L5, R2, R3, R4 | Not assessed | 0 | 0 | None | None | None |
| SITE2_022 | PsA | 1 | 30s | Female | 6 | L1, L3, L4, L5, R1, R3, R4 | None, L1, L2, L3, L4, L5, R1, R2, R3, R4, R5 | 0 | 0 | None | None | None |
| SITE2_023 | PsO | 1 | 50s | Male | 11 | L2 | None | 0 | 0 | None | None | None |
| SITE2_024 | PsA | 1 | 30s | Male | 7 | L1, L2, L3, L4, L5, R1, R2, R3, R4, R5 | L1, L2, L3, L4, L5, R1, R2, R3, R4, R5 | 7 | 5 | None | R3 | L2, L4, R2 |
| SITE2_025 | PsA | 1 | 40s | Male | 6 | None | None | 7 | 6 | None | None | None |
| SITE2_030 | PsA | 1 | 50s | Male | 0.4 | None | None | 3 | 3 | right_achilles | None | None |
| SITE2_031 | PsA | 1 | 30s | Female | 10 | None | None | 17 | 5 | None | None | L4, R2 |
| SITE2_032 | PsO | 1 | 50s | Male | 5 | L1, L2, L5, R1, R3, R4 | None | 0 | 0 | None | None | None |
| SITE2_033 | PsA | 1 | 50s | Female | 2 | None | None | 7 | 7 | right_arm, left_arm, right_achilles, left_achilles | None | None |
| subject 03 | PsO | 1 | 20s | Male | 15 | None | None | 0 | 0 | None | None | None |
| subject 10 | PsA | 1 | 40s | Male | 0 | None | None | 0 | 0 | None | None | None |
| subject 12 | PsA | 1 | 60s | Male | 0 | None | None | 0 | 0 | None | None | R3, R4 |
| subject 13 | PsO | 1 | 20s | Male | 3 | None | None | 0 | 0 | None | None | None |
| subject 16 | PsA | 1 | 30s | Female | 5 | None | None | 0 | 0 | None | None | None |
| subject 17 | PsA | 1 | 40s | Male | 0 | None | None | 0 | 0 | None | None | None |
| subject 19 | PsA | 1 | 40s | Male | 0 | None | None | 0 | 0 | None | None | None |
| subject 20 | PsA | 1 | 20s | Female | 0.5 | None | L1, R1 | 0 | 0 | None | None | None |
| subject 22 | PsA | 1 | 40s | Male | 0 | None | None | 0 | 0 | None | None | R3 |
| subject 24 | PsA | 1 | 20s | Female | 1 | None | None | 0 | 0 | None | None | R3 |
| subject 27 | PsA | 1 | 60s | Female | 0 | None | None | 0 | 0 | None | None | None |
| subject 30 | PsA | 1 | 50s | Female | 0 | None | None | 0 | 0 | None | None | L2, L3, L4 |
| subject 31 | PsA | 1 | 40s | Male | 0 | None | None | 0 | 0 | None | None | None |
| subject 32 | PsA | 1 | 30s | Male | 0 | None | None | 0 | 0 | None | None | None |
| subject 33 | PsA | 1 | 30s | Male | 0 | None | None | 0 | 0 | None | None | None |
| subject 36 | PsA | 1 | 30s | Male | 0 | None | None | 0 | 0 | None | None | None |
| subject 37 | PsA | 1 | 50s | Female | 0 | L4 | None | 4 | 2 | None | None | None |
| subject 50 | PsA | 1 | 30s | Male | 1.5 | None | None | 0 | 0 | None | None | None |
| subject 51 | PsA | 1 | 40s | Male | 0 | L1, L2, L3, L4, L5, R1, R2, R3, R4, R5 | L1, R1 | 14 | 12 | None | L2, L3 | L4, R1 |
| subject 55 | PsA | 1 | 50s | Male | 1 | Not assessed | Not assessed | 0 | 0 | left_arm, right_arm | None | None |
| subject 56 | PsA | 1 | 50s | Male | 2 | Not assessed | None | 0 | 0 | None | R2 | L4, R5 |
| subject 60 | PsA | 1 | 30s | Female | 0 | None | None | 0 | 0 | left_arm, right_arm | None | None |
| SITE1_008 | PsA | 2 | 30s | Male | 0.5 | L4 | L1, L2, L3, L4, L5, R1, R2, R3, R4, R5 | 0 | 0 | None | None | None |
| SITE1_022 | PsA | 2 | 50s | Male | 0 | None | None | 0 | 0 | None | None | None |
| SITE1_030 | PsA | 2 | 20s | Male | 0.5 | L1, L2, L3, L4, L5, R1, R2, R3, R4, R5 | L1, L2, L3, L4, L5, R1, R2, R3, R4, R5 | 0 | 0 | None | None | R3, R4 |
| SITE1_037 | PsA | 2 | 50s | Female | 0 | None | L1, L2, L3, L4, L5, R1, R2, R3, R4, R5 | 0 | 0 | None | None | None |
| SITE2_006 | PsO | 2 | 20s | Female | 13 | None | None | 0 | 0 | None | None | None |

| | | | | | | | | | | | | |
|---|---|---|---|---|---|---|---|---|---|---|---|---|
| SITE2_009 | PsA | 2 | 40s | Female | 10 | None | None | 0 | 0 | left_arm, right_arm, right_achilles | None | None |
| subject 24 | PsA | 2 | 20s | Female | 0.5 | None | None | 0 | 0 | None | None | R3 |
| subject 37 | PsA | 2 | 50s | Female | 0.5 | Not assessed | None | 0 | 0 | None | None | None |
| subject 50 | PsA | 2 | 30s | Male | 0.5 | L1 | None | 0 | 0 | None | None | None |
| SITE2_006 | PsO | 3 | 30s | Female | 1 | None | None | 0 | 0 | None | None | None |
| subject 24 | PsA | 3 | 20s | Female | 0.5 | None | None | 0 | 0 | None | None | L3 |
| Subject 37 | PsA | 3 | 50s | Female | 0 | None | None | 0 | 0 | right_knee, left_knee | None | None |

### Confounding analyses supplement

#### 1 Motivation

As described in the main text, one of the objectives of our study was to evaluate if the total-rotation (overall rotation) feature derived from the Digital Jar Open activity could be used for: (i) training classifiers of upper extremity involvement (UEI) using records collected from controls, people with psoriatic arthritis, and people with psoriasis; and (ii) training classifiers of psoriatic arthritis diagnosis without upper extremity involvement (PsA-without-UEI), that is, classifiers of PSA vs psoriasis (PsO), using only a subset of the individuals with PsA that do not show UEI[1]. Figure S1 reports the AUROC scores for classifiers trained and evaluated in 1000 random splits of the data, and show that they can achieve a median AUROC score of 0.72 and 0.76 on these two classification tasks. Similarly, Figure S2 reports the AUPRC scores.

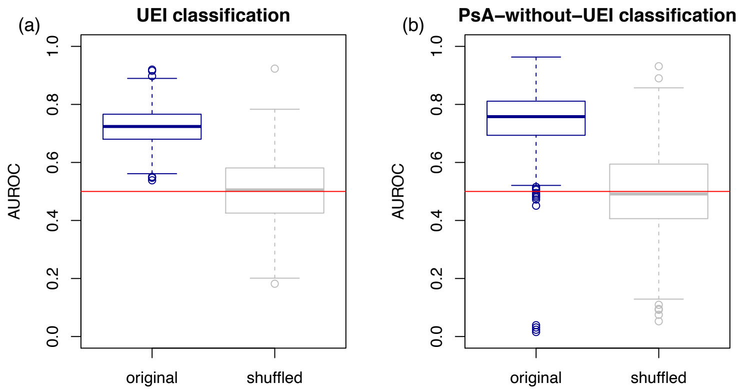


**Figure S1: Unadjusted classifier performances based on AUROC.** Panel a shows the results for the UEI classifier, while panel b reports the results for the PsA-without-UEI classifier. In both panels, the blue boxplots show the distribution of the AUROC scores computed on 1000 distinct random splits of the data into training and test sets. The grey boxplots report the results for classifiers trained and evaluated on shuffled labels, and provide information about the range of AUROC scores we would expect to see by chance for our test set sizes.

[1]These individuals might include: people with pre-clinical damage to their arm joints/entheses that is not detectable by a physician (but may be detectable by power doppler/ultrasound techniques); people with painful or swollen joints outside of their upper extremities (e.g. legs, fingers, toes); and people with swollen, but not painful, shoulders, elbows, or wrists.

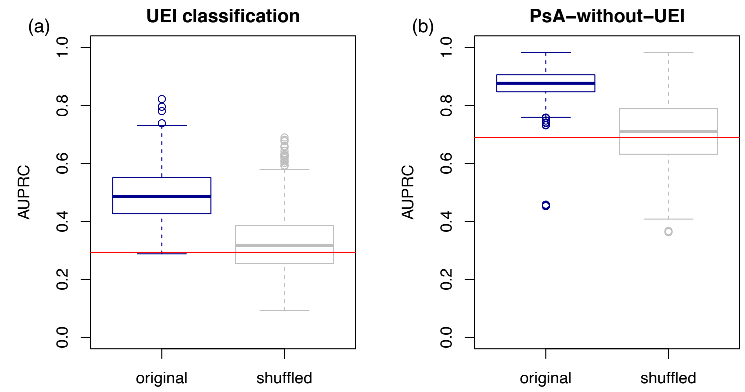


**Figure S2: Unadjusted classifier performances based on AUPRC.** Panel a shows the results for the UEI classifier, while panel b reports the results for the PsA-without-UEI classifier. The red lines represent the random guess baseline performance, which corresponds to the proportion of positive cases in the data. (For the UEI classifier we have 22 positive (pain) and 53 negative cases (no pain), so that the baseline is approximately 0.29. For the PsA-without-UEI classifier we have 31 positive (PsA) and 14 negative (PsO) examples, so that the baseline is given by 0.69.) In both panels, the blue boxplots show the distribution of the AUPRC scores computed on 1000 distinct random splits of the data into training and test sets. The grey boxplots report the results for classifiers trained and evaluated on shuffled labels, and provide information about the range of AUROC scores we would expect to see by chance for our test set sizes.

However, as illustrated in Figures S3 a and b, the total-rotation input is negatively associated with age (so that records from older individuals tend to achieve lower total-rotation scores). Similarly, Figures S3 c and d report weak associations between age and the two pain labels (showing that records from older individuals tend to report the presence of pain more frequently than the records from younger individuals).

These observations raise the possibility that the predictive ability of our classifiers might be due to the fact that total-rotation might be a good feature to tell apart young from old individuals, rather than to classify UEI or PsA-without-UEI (and the only reason we observe a positive predictive ability is because age is associated with the classifiers labels). In other words, age might be confounding our classifiers' predictions.

In machine learning applications, the presence of confounding can lead to ambiguous inference and poor generalizability of models in situations where selection mechanisms generate dataset shifts [2] between the training and test sets. For instance, consider a (hypothetical) scenario where our classifiers are mostly picking up the age signal, and where they are going to be deployed in a population where the association between labels

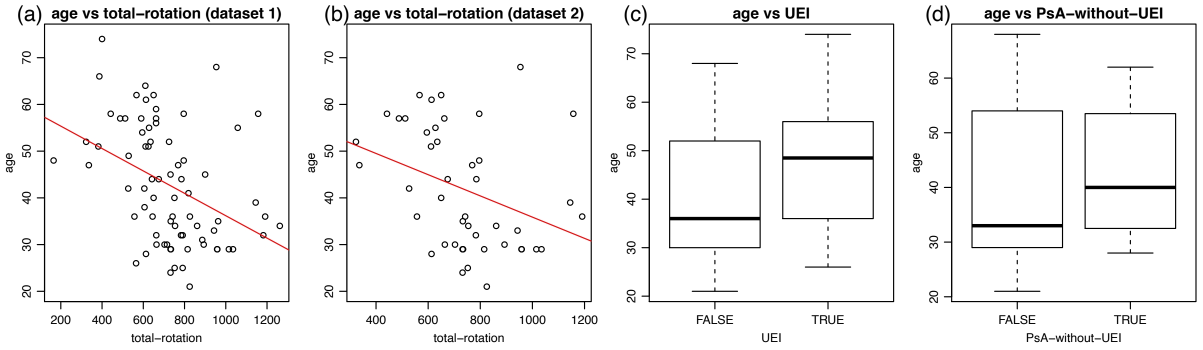


**Figure S3: Age as a potential confounder.** Panels a shows a negative association between total-rotation and age in the dataset (dataset 1) used for building the UEI classifier, while panel b shows the analogous result for the smaller subset (dataset 2) used for building the PsA-without-UEI classifier. Panels b and c show, respectively, weak associations between age and UEI the between age and PsA-without-UEI labels.

and age are different from the training set (e.g., where younger rather than older individuals tend to report more UEI). In this scenario, such classifiers would likely perform quite poorly, as they would tend to predict that older individuals show UEI. Confounding is an important practical issue because dataset shifts affecting the joint distribution of confounders and outcomes are quite common in real word applications, where selection biases [3, 4] often lead to the collection of non-representative training sets. Ideally, we would want to build classifiers that are unable to pick up confounding signals and, therefore, are better able to generate stable predictions that are more robust to dataset shifts generated by selection biases.

Here, we describe approaches to evaluate if observed demographic/metadata variables (namely, age, gender, and study site) are indeed confounding our predictions, as well as, an approach to adjust model inputs and generate unconfounded and stable predictions.

The remaining of this supplement is organized as follows:

- Section 2 ("Background") defines the notation and provides key causal inference definitions and concepts necessary to understand the analyses in this supplement. (Confounding is a causal concept which cannot be described in terms of associations alone, and requires the use of causal diagrams describing our qualitative assumptions about the causal relations between the variables [5]. See also chapter 4 of reference [7] for a less technical introduction to this key concept.) This self contained section is included to help readers unfamiliar with causal inference methods.

- Section 3 ("Informal first checks") describes some informal analyses that can be used to discard some potential confounders, without the need for more careful analyses. It only involves building ML models for predicting the confounders (and do not require causal inference techniques). The basic idea is that if our input (total-rotation) is not able to predict the confounders, then the input does not capture the confounder signals and do not need any adjustments.

- Section 4 ("Confounding evaluation") uses the causal inference methodology described in [9] to evaluate whether the variables that passed the initial informal checks described in Section 3 are indeed confounding our predictions.
- Finally, Section 5 ("Confounding adjustment") describes the causality-inspired approach proposed in [11] for generating unconfounded and stable predictions, and illustrate its application to deconfound our predictions.

### 2 Background

Throughout this supplement we let $X$ represent the input variable (i.e., the feature), $D$ represent the disease status label (i.e., psoriatic arthritis, psoriasis, or control), $Y$ represent the UEI label, and $C$ represent a putative confounder variable. (Note that we use the "putative" qualifier to make clear that a variable $C$ might or might not turn out to be a confounder.)

We adopt the graph-based approach to causality championed by Pearl [5], where the statistical associations encoded in the joint probability distribution of a set of random variables is supplemented by a directed acyclic graph (DAG) describing the causal relations between the variables. In a causal DAG, a directed edge (arrow) $A \rightarrow B$ indicates that $A$ is a cause of $B$.

In this document we will only deal with "anticausal prediction tasks" [6], which correspond to prediction tasks where the prediction goes in the opposite direction of the causal relation between the outcome and the inputs, that is, where the outcome is a cause of the inputs. ("Causal prediction tasks", on the other hand, correspond to prediction tasks where the inputs represent the causes of the outcome.)

Under Pearl's graph-based approach to causality, a variable $C$ is a confounder of the relationship between a feature, $X$, and the labels, $Y$, if there is a path from $C$ to $X$ that does not go thorough $Y$, and there is a path from $C$ to $Y$ that does not go thorough $X$. (Where a path is defined as any unbroken, non-intersecting sequence of directed edges, which may go along or against the direction of the arrows.) As an example, consider the causal diagrams $M_1$, $M_2$, $M_3$, and $M_4$ presented in Figure S4. The variable $C$ is a confounder only on diagram $M_1$, since the path $C \rightarrow X$ connects $C$ to $X$ without going through $Y$, and the path $C \rightarrow Y$ connects $C$ to $Y$ without going through $X$. For diagram $M_2$, the only path connecting $C$ to $X$ goes thorough $Y$. For diagram $M_3$, the only path connecting $C$ to $Y$ goes thorough $X$. For diagram $M_4$ there are no paths connecting $C$ to $X$ or $C$ to $Y$.

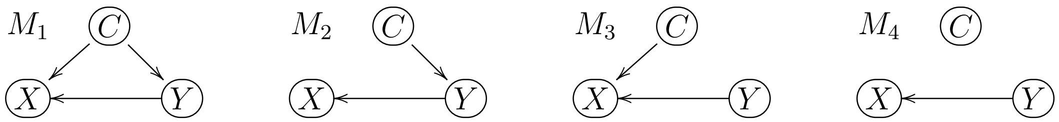


**Figure S4:** The variable $C$ satisfies the definition of a confounder only in model $M_1$.

Intuitively, we have that $C$ is a confounder only on model $M_1$ because only in this model the total association between $X$ and $Y$ is given by the combination of: (i) the "direct association" generated by the causal effect of $Y$ on $X$ (through the path $Y \rightarrow X$); and (ii) the "spurious association" generated by the confounder $C$ (through the path $X \leftarrow C \rightarrow Y$). For all other models, the association between $X$ and $Y$ is generated by the causal effect of $Y$ on $X$ alone.

As described in detail in Section 4, the inspection of marginal and conditional associations in the data play a key role for determining whether $C$ is really a confounder. We adopt the notation $\perp\!\!\!\perp$ and $\not\!\perp\!\!\!\perp$ to describe statistical independence and dependence, respectively, where $Z_1 \perp\!\!\!\perp Z_2$ indicates that $Z_1$ and $Z_2$ are marginally independent, while $Z_1 \not\!\perp\!\!\!\perp Z_2$ indicates marginal dependency. Similarly, we use the notation $Z_1 \perp\!\!\!\perp Z_2 \mid Z_3$, to describe that $Z_1$ is independent of $Z_2$ conditional on $Z_3$, and $Z_1 \not\!\perp\!\!\!\perp Z_2 \mid Z_3$, to describe that $Z_1$ and $Z_2$ are still associated conditional on $Z_3$.

An important tool that will be used to determine if $C$ is a confounder is the concept of d-separation proposed by [5]. In a nutshell, d-separation provides a set of simple graphical rules that allows the determination the conditional independence relationships between the variables in a causal diagram. (In other words, the d-separation rules allow us to "read out" all the marginal and conditional (in)dependence relationships among the variables in the diagram by simply inspecting the diagram. Basically, when the diagram shows that a given variable, say $Z_3$, "blocks" all paths between variables $Z_1$ and $Z_2$, we have the $Z_1$ is independent of $Z_2$ conditional on $Z_3$.) Explicitly, the d-separation rules are given by:

*d-separation* [5]: A path is *d-separated* or *blocked* by a set of nodes $\boldsymbol{W}$ if and only if:

1. The path contains a chain $Z_j \rightarrow Z_m \rightarrow Z_k$ such that the middle node $Z_m$ is in $\boldsymbol{W}$.
2. The path contains a fork $Z_j \leftarrow Z_m \rightarrow Z_k$ such that the middle node $Z_m$ is in $\boldsymbol{W}$.
3. The path contains a collider $Z_j \rightarrow Z_m \leftarrow Z_k$ such that $Z_m$ is not in $\boldsymbol{W}$ and no descendant of $Z_m$ is in $\boldsymbol{W}$.

We say that a path is d-connected or open when it is not d-separated or blocked. We also say that a joint probability distribution over a set of variables is faithful [5, 8] to a DAG representing the causal relationships between these variables if no conditional independence relations, other than the ones implied by the d-separation criterion are present.

Intuitively, it is useful to think about the d-separation as the rules that govern the flow of information between the variables in the diagram. When a path is closed or d-separated, the information flow is blocked, whereas when it is open (or d-connected) information is still allowed to flow. As an example, the application of d-separation to model $M_2$ in Figure S4 shows that,

$$X \not\perp\!\!\!\perp Y \; , \;\; X \not\perp\!\!\!\perp C \; , \;\; C \not\perp\!\!\!\perp Y \; , \;\; X \not\perp\!\!\!\perp Y \mid C \; , \;\; X \perp\!\!\!\perp C \mid Y \; ,$$

where we see that $X$ is independent of $C$ given $Y$ (i.e., $X \perp\!\!\!\perp C \mid Y$) since, if we already know the value of $Y$, then $C$ no longer have any additional information about $X$.

### 3 Informal first checks

Our first step was to check whether the total-rotation feature captures age, gender, or site related signals. To this end we trained machine learning (ML) models for predicting the confounder variables, using the total-rotation as the input of the ML model. We adopt a linear regression model for predicting age and logistic regression models for classifying gender and site. Figure S5 report the results based on 1000 random splits of the data into training and test sets.

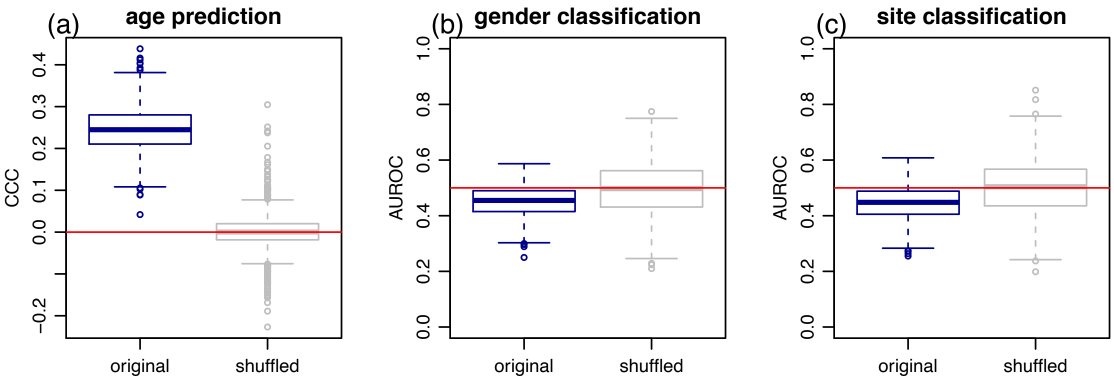


**Figure S5: Confounder predictions.**

Figure S5 a clearly shows that total-rotation can be used to predict age. Note how the predictive performance on the original data (blue boxplot), measured by the concordance correlation coefficient (CCC), is clearly above the random guess baseline performance (grey boxplot centered at zero) obtained by regression models trained on shuffled outcome data (i.e., where we randomly shuffled the age values before training and evaluating the regression models). Figures S5 b and c, on the other hand, show that total-rotation is unable to predict gender or site. (Note that the predictive performance for these classifiers is slightly worse than a random guess baseline performance, centered at an AUROC value of 0.5, obtained by classifiers trained on shuffled labels. However, as reported in Figure S6, these slightly worse than random performances are not statistically significant.)

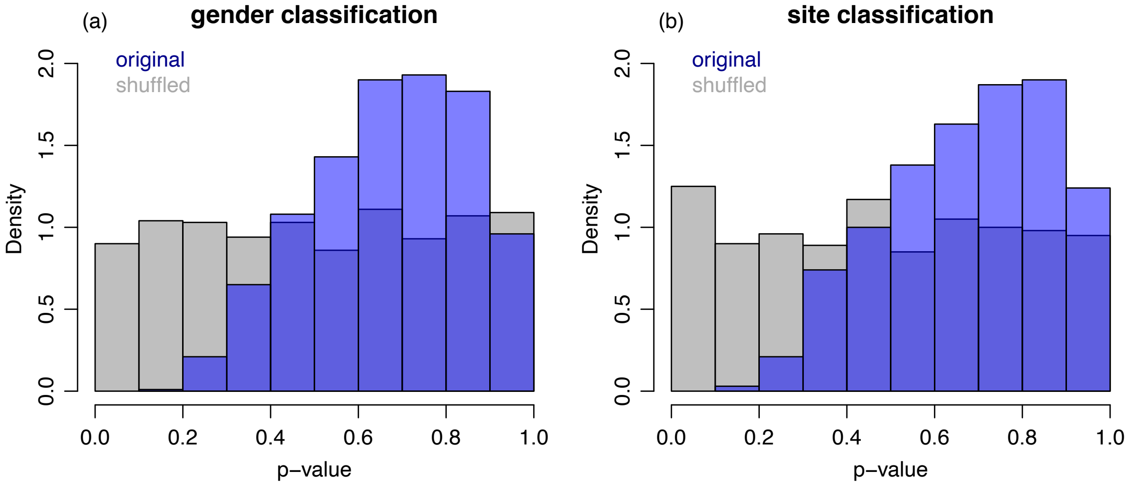


**Figure S6: P-value distributions for the gender (panel a) and site (panel b) classifiers.** P-values from a statistical test for checking if the classification performance was better than a random guess. (Computed from a transformation of the one tailed Mann-Whitney test comparing the null hypothesis that $AUROC = 0.5$ against the alternative that $AUROC > 0.5$, as described in [1].) Grey and blue histograms report the results based on classifiers built with shuffled and original labels, respectively, and show that the classification was not statistically better than a random guess. Results based on the 1000 data splits. On a technical note, this analytical test assumes that the data points are independent, what is not strictly true in our data (since we have a reduced number of repeated measurements (2 or 3 records) on 3 individuals). Nonetheless, repeating the analysis after removing the repeated measurements still produce similar results.

Note, as well, that these results are consistent with the fact that total-rotation is (marginally) associated with age but not with gender or site as shown in Figure S7.

These results suggest that neither gender nor site appear to be confounding the predictive performance of our classifiers, as the total-rotation feature does not seen to be able to capture the gender or the site signals. Age, on the other hand, might potentially be confounding our results (although the more careful additional analyses described in the next section are needed to show this is indeed the case).

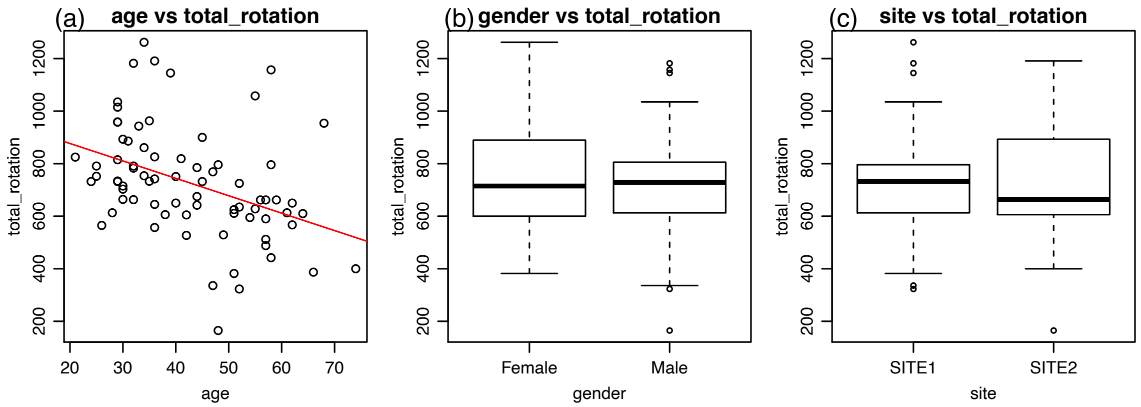


**Figure S7: Confounders vs total-rotation associations.** Panel a reports a (statistically significant) negative correlation between age and total-rotation ($\hat{\rho} = -0.4$, $p = 3.96 \times 10^{-4}$), showing that older individuals tend to achieve smaller total-rotations than younger ones. Panels b and c report, respectively, weak (and not significant) associations between gender and total-rotation ($\hat{\rho} = -0.05$, $p = 0.697$) and between site and total-rotation ($\hat{\rho} = 0.03$, $p = 0.808$). (For the correlation tests with the categorical variables gender and site, we first converted the levels to binary variables. Namely, the "Female" and "Male" levels of gender and the "Site 1" and "Site 2" levels of the site variables were coded as 0 and 1, respectively.)

### 4 Confounding evaluations

As described in detail in reference [9] we can determine if age is indeed confounding our UEI classifier predictions by checking if the conditional independence (CI) relations among the total-rotation, the UEI label, and the age variables observed in the test set data match the CI relations implied by the application of d-separation to the causal diagram representing how the data was generated. (Similarly, for the PsA-without-UEI classifier, we can check the CI relationships between total-rotation, the disease label, and age.)

Hence, the first step in a confounding analysis is to draw a causal diagram describing how the input, outcome, and potential confounder variables are related. In our application, the data generation process underlying the data can be described by causal diagrams such as in Figure S8, where $D$ represents a categorical variable reporting the disease status (psoriatic arthritis, psoriasis, and controls), $Y$ represents a binary variable reporting the UEI label, $X$ represents the total-rotation, and $C$ represents a putative confounder such as age, gender, or site. Note that, in all diagrams, we have that $D \rightarrow X$, since the disease status can certainly affect the total-rotation measurement (since people with compromised joints, due to psoriatic arthritis, are expected to show more constrained rotation ability than controls or individuals with psoriasis but no psoriatic arthritis), while

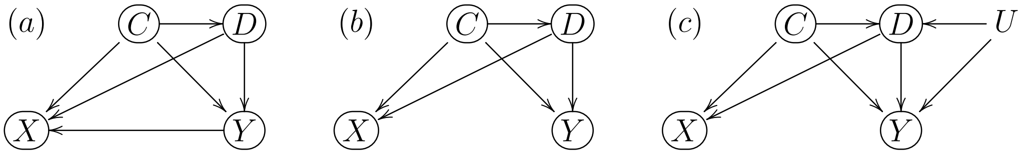


**Figure S8: Possible causal diagrams underlying the observed data.**

someone's ability to rotate a smartphone over a table cannot cause the disease. (In other words, reduced total-rotation measurements represent a symptom of the disease.) Similarly, we have that $D \to Y$ since the joint inflammation characterizing psoriatic arthritis can generate UEI, but not the other way around. (Again, pain is a symptom of the inflammation process and not its cause. This relationship might, nonetheless, be confounded by additional unmeasured variables $U$ that might cause psoriatic inflammation and pain, as described in Figure S8 c.)

We also see in all diagrams that $C \to X$ since potential confounders such as age, gender, or site can impact the total-rotation, while total-rotation cannot be a cause of these variables. Observe, as well, that in all diagrams we have that $C \to D$ and $C \to Y$, since potential confounders such as age and gender might also influence $D$ and $Y$ (since age is a risk factor for psoriatic arthritis and older age is also a cause for UEI, whereas $D$ and $Y$ are certainly not causes of age)[2]. Finally, regarding the causal relationship between $Y$ and $X$, while it is possible that $Y$ has a direct causal effect on $X$ as depicted in Figure S8 a (since feeling pain might preclude someone from rotating the smartphone well), it might also be the case that $Y$ does not directly affect $X$ and that the association between $Y$ and $X$ is generated by their common causes $C$ and $D$, as depicted in Figures S8 b and c.

For the remaining of this section we will focus on the diagram in Figure S8 a (although adopting the other diagrams would also work for detecting confounders using d-separation).

For our analysis, rather than working directly with the "data generation process" causal diagram (Figure S8 a) we will consider the "classification task" causal diagrams, which augment the data generation causal diagram with the "prediction generation process" (as this allow us to directly assess whether the classifier predictions are confounded[3]). The augmented diagrams for the UEI and PsA-without-UEI classification tasks are presented, respectively, in Figure S9 a and b,

[2]We point out, however, that for other potential confounders such as site, the association between the $D$ and $C$ and between $Y$ and $C$ might be generated by selection mechanisms rather than by an effect of the confounder on the disease status or pain label. This difference is, nonetheless, inconsequential for our analyses, as it does not impact the conditional independence relationships between $C$, $D$, $Y$, and $X$.

[3]Also, when our ML models contain multiple inputs (what is not the case for our analyses, but is usually the case in most applications) it is more convenient to work with the prediction $\hat{R}_{ts}$ directly, rather than with multiple inputs.

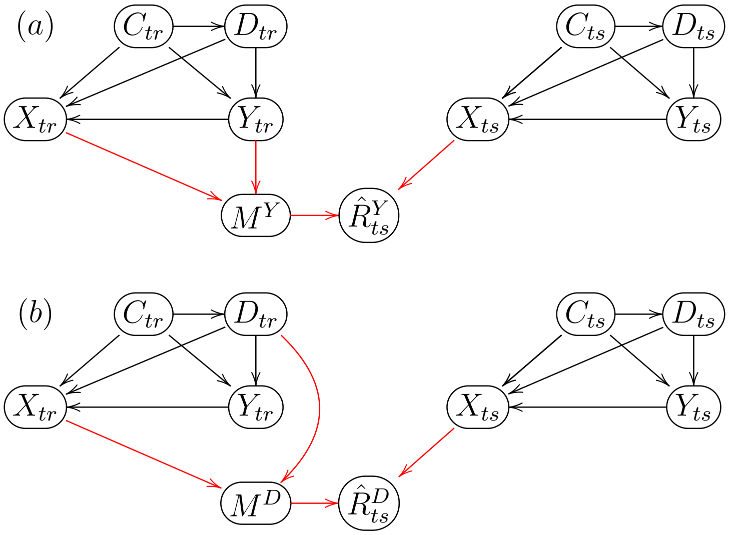


**Figure S9: Confounded classification tasks.** Panel a shows the full "classification task" diagram for the UEI classification task, while panel b shows the respective diagram for the PsA-without-UEI task. In both panels, the black arrows represent the "data generation process", while the red arrows represent the "prediction generation process".

where the subscripts $tr$ and $ts$ indicate the training and test sets, respectively, $M^Y$ and $M^D$ represent the trained classification models, and $\hat{R}^Y_{ts}$ and $\hat{R}^D_{ts}$ represent the predicted positive class probability generated by the classifier. Note that $\hat{R}^Y_{ts}$ is a function of the training data (i.e., training labels, $Y_{tr}$, and training features, $X_{tr}$), as well as, the test features, $X_{ts}$, since $\hat{R}^Y_{ts}$ is generated by evaluating the test features using the model trained on the training data. (Similarly, $\hat{R}^D_{ts}$ is a function of $D_{tr}$, $X_{tr}$, and $X_{ts}$.) Figures S10 and S11 present, respectively, simplified versions of the diagrams in Figure S9 a and b, focusing only on the variables of interest for each classification task.

For the UEI classification task, by focusing on the simplified diagram in Figure S10 b (see figure caption for further details) we have that the CI relations implied by the application of the d-separation criterion to the diagram are given (under the faithfulness assumption) by,

$$\hat{R}^Y_{ts} \not\perp\!\!\!\perp Y_{ts} \ , \ \ \hat{R}^Y_{ts} \not\perp\!\!\!\perp C_{ts} \ , \ \ C_{ts} \not\perp\!\!\!\perp Y_{ts} \ , \ \ \hat{R}^Y_{ts} \not\perp\!\!\!\perp Y_{ts} \mid C_{ts} \ , \ \ \hat{R}^Y_{ts} \not\perp\!\!\!\perp C_{ts} \mid Y_{ts} \ . \tag{1}$$

Similarly, for the PsA-without-UEI classifier, the CI relations implied by d-separation are,

$$\hat{R}^D_{ts} \not\perp\!\!\!\perp D_{ts} \ , \ \ \hat{R}^D_{ts} \not\perp\!\!\!\perp C_{ts} \ , \ \ C_{ts} \not\perp\!\!\!\perp D_{ts} \ , \ \ \hat{R}^D_{ts} \not\perp\!\!\!\perp D_{ts} \mid C_{ts} \ , \ \ \hat{R}^D_{ts} \not\perp\!\!\!\perp C_{ts} \mid D_{ts} \ . \tag{2}$$

Hence, in order to evaluate if age is indeed confounding our UEI predictions, $\hat{R}^Y_{ts}$, we need to check if the CI relations in (1) hold in the data. (Similarly, for the PsA-without-UEI classifier we need to check if the CI relations in (2) hold in the data.) Figure

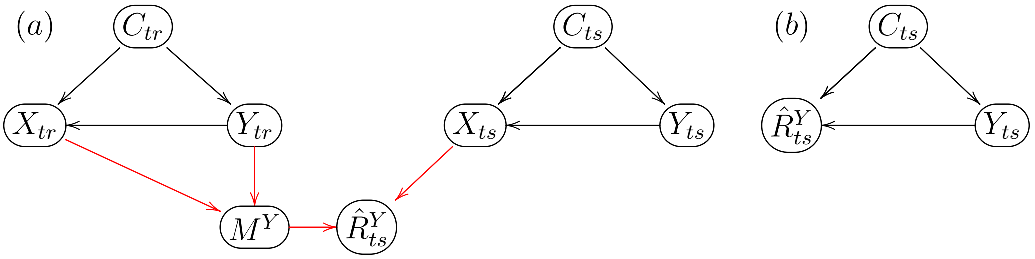


**Figure S10: Simplified confounded classification task for the UEI outcome.** Note that by ignoring the $D$ variable, we have in panel a that the arrow $C \rightarrow X$ actually represents the combined paths $C \rightarrow X$ and $C \rightarrow D \rightarrow X$ in Figure S9 a, while the arrow $C \rightarrow Y$ actually represents the combined paths $C \rightarrow Y$ and $C \rightarrow D \rightarrow Y$ in Figure S9 a. Panel b shows a simplified version focusing only on the prediction scores, test set labels and confounders, where the path $C_{ts} \rightarrow X_{ts} \rightarrow \hat{R}^Y_{ts}$ is replaced by the collapsed version $C_{ts} \rightarrow \hat{R}^Y_{ts}$, and the paths $Y_{ts} \rightarrow X_{ts} \rightarrow \hat{R}^Y_{ts}$ is replaced by the path $Y_{ts} \rightarrow \hat{R}^Y_{ts}$.

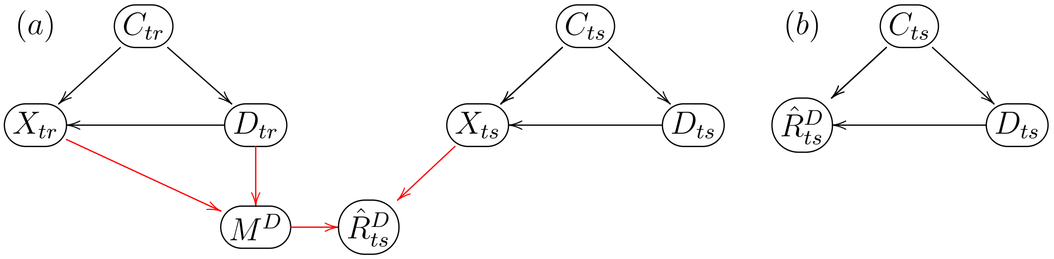


**Figure S11: Simplified confounded classification task for the PsA-without-UEI classifier.** Note that by ignoring the $Y$ variable, we have in panel a that the arrow $C \rightarrow X$ actually represents the combined paths $C \rightarrow X$ and $C \rightarrow Y \rightarrow X$ in Figure S9 b, while the arrow $D \rightarrow X$ actually represents the combined paths $D \rightarrow X$ and $D \rightarrow Y \rightarrow X$ in Figure S9 b. Panel b shows a simplified version focusing only on the prediction scores, test set labels and confounders, where the path $C_{ts} \rightarrow X_{ts} \rightarrow \hat{R}^D_{ts}$ is replaced by the collapsed version $C_{ts} \rightarrow \hat{R}^D_{ts}$, and the paths $D_{ts} \rightarrow X_{ts} \rightarrow \hat{R}^D_{ts}$ is replaced by the path $D_{ts} \rightarrow \hat{R}^D_{ts}$.

S12 a reports the distributions (calculated over the 1000 data splits) of the estimated correlations and partial correlations among the $\hat{R}_{ts}$, $Y_{ts}$, and $C_{ts}$ variables for the UEI classifiers, while Figure S12 b reports the analogous quantities for the PsA-without-UEI classifier. The results show that the observed marginal and conditional associations are consistent with the CI relations in (1) and (2), showing that age is indeed confounding the classifier predictions.

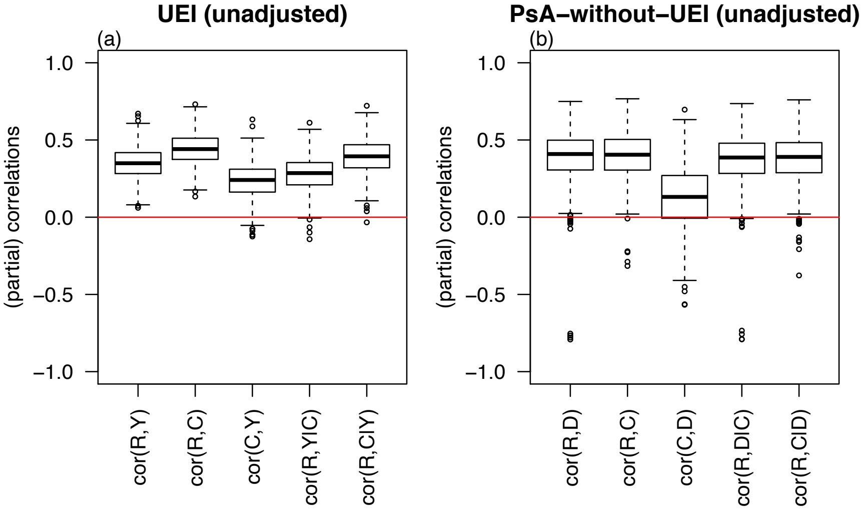


**Figure S12: CI pattern evaluations for classifiers trained with the original (unadjusted) total-rotation input.** $R$ represents the test set prediction $\hat{R}_{ts}^{Y}$ or $\hat{R}_{ts}^{D}$, $C$ represents the age, $Y$ represents the UEI label, and $D$ presents the disease status, namely, PsA (without UEI) or PsO.

### 5 Confounding adjustment

A popular approach for deconfounding predictions in ML applications is to balance the labels and confounder variables using matching techniques before training and evaluating the classifiers. Unfortunately, the small sample size of our study makes it unfeasible to perform matching in our data. (Other balancing techniques such as oversampling from the minority classes are also problematic in small datasets due to the increased risk of overfitting, and more sophisticated balancing techniques that use synthetic data to balance the dataset such as SMOTE [10] cannot be applied with a single input variable.)

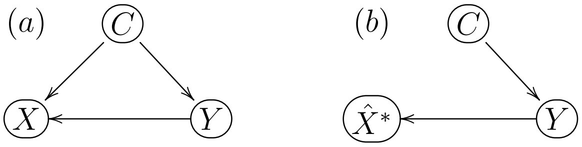


**Figure S13: Causality-aware adjustment.** Panel a shows the confounded model where the confounder, $C$, directly affects the input, $X$. Panel b shows the adjusted model where the adjusted input, $\hat{X}^*$, is no longer directly influenced by the confounder. (Note that for the PsA-without-UEI classifier, the adjustment is conducted by replacing the label $Y$ by the label $D$.)

Here, we adopt the causality-aware adjustment proposed in [11]. As illustrated in Figure S13, the basic idea is to adjust the ML model inputs (by removing the confounding signal) and then build ML models using the adjusted inputs, which retain only the associations generated by the causal influence of the label on the features.

The approach assumes that the input variables are generated according to linear structural causal models and, in the case of a single feature and a single confounder, it is implemented as follows (see [11] for the general case):

**Causality-aware adjustment:**

1. Using the training set, estimate regression coefficients and residuals from the linear model,
$$X_{tr} = \mu_X^{tr} + \beta_{XY}^{tr}\, Y_{tr} + \beta_{XC}^{tr}\, C_{tr} + W_X^{tr} \ ,$$
and then compute the respective adjusted feature as,
$$\hat{X}_{tr}^{*} = \hat{\mu}_X^{tr} + \hat{\beta}_{XY}^{tr}\, Y_{tr} + \hat{W}_X^{tr} = X_{tr} - \hat{\beta}_{XC}^{tr}\, C_{tr} \ .$$

2. Using the test set, compute the adjusted feature,
$$\hat{X}_{ts}^{*} = X_{ts} - \hat{\beta}_{XC}^{tr}\, C_{ts} \ ,$$
using the regression coefficient, $\hat{\beta}_{XC}^{tr}$, estimated in the training data.

One advantage of the approach over the balancing adjustments is that it is able to generate stable predictions that are robust to dataset shifts in the association between the labels and confounders generated by selection biases [11, 12]. One disadvantage is that, contrary to the balancing methods which are non-parametric adjustment approaches, the causality-aware adjustment makes the assumption that a linear model provides a good fit to the data (and might fail to deconfound the predictions when this is not the case). Hence, it is essential to evaluate the adjustment performance using the CI pattern approach described in the previous section to make sure the adjustment is working.

Figure S14 reports the AUROC scores for classifiers trained with the age-adjusted total-rotation input (green boxplots). (Figure S15 reports the distribution of the p-values for testing whether $AUROC > 0.5$ and indicates that even after adjustment the classifiers are performing better than a random guess.) Comparison with the scores generated by classifiers trained with the unadjusted total-rotation (blue boxplots in Figure S14) shows a small decrease in predictive performance, indicating that age contributes to the predictive performance to some extent. (Observe, however, that because we are randomly splitting the data into training and test sets, we prevent dataset shifts. However, in practice, if we deploy these classifiers in different test sets where the association between age and the labels is different from the association in the training set, we will likely see an unstable performance for the unadjusted classifiers, while the adjusted ones should be more stable.) Similarly, S16 reports analogous results for the AUPRC metric.

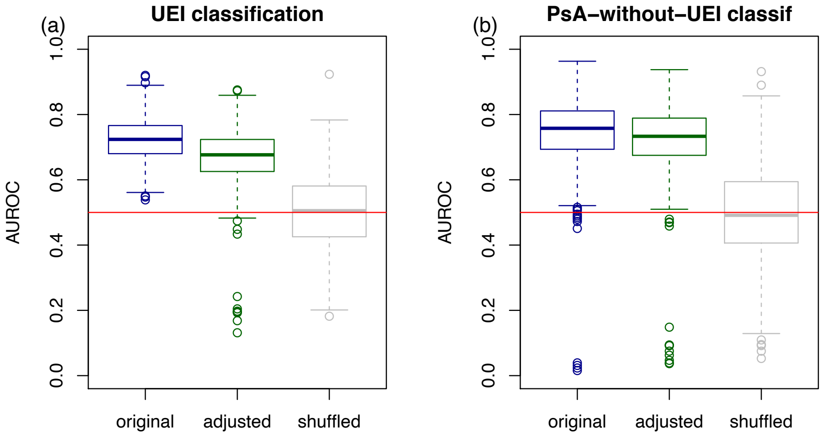


**Figure S14: Unadjusted vs adjusted classifier performances according to the AUROC metric.** The blue and green boxplots report the distribution of the AUROC scores from classifiers build with unadjusted and age-adjusted inputs. As before, the grey boxplot report the results based on shuffled labels.

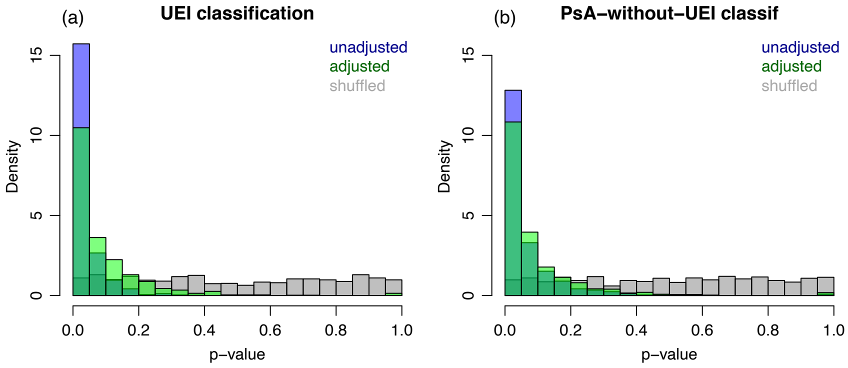


**Figure S15: P-value distributions for the UEI and PsA-without-UEI classifiers.** P-values from a statistical test for checking if the classification performance was better than a random guess. (Computed from a transformation of the one tailed Mann-Whitney test comparing the null hypothesis that $AUROC = 0.5$ against the alternative that $AUROC > 0.5$, as described in [1].) Grey, green, and blue histograms report the results based on classifiers built with shuffled labels, adjusted data, and original data, respectively. Results based on the 1000 data splits.

As mentioned above, because the causality-aware adjustment assumes a linear model for the input variables, it is important to check if the adjustment is working as expected.

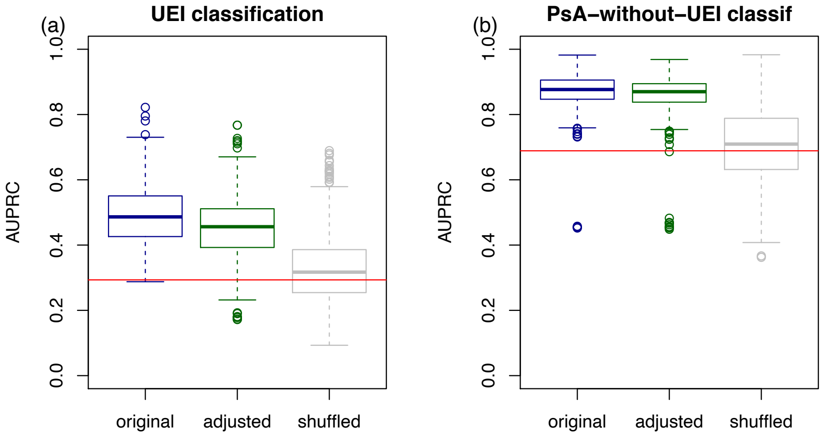


**Figure S16: Unadjusted vs adjusted classifier performances according to the AUPRC metric.** The blue and green boxplots report the distribution of the AUPRC scores from classifiers build with unadjusted and age-adjusted inputs. As before, the grey boxplot report the results based on shuffled labels.

To this end, we repeated the analyses presented in Sections 3 and 4 for classifiers trained with the age-adjusted total-rotation input. Figure S17 shows that the adjusted total-rotation (green boxplot) no longer is able to predict age. Figure S18 shows that the conditional independence patterns between the test set labels, age, and the classifier predictions are no longer consistent with the CI relations we would expect to see in a confounded classifier. For instance, for the UEI classifier (Figure S18 a) note how the distribution of $cor(C, R \mid Y)$ is centered around zero, indicating that age, $C_{ts}$, is conditionally independent of the prediction, $\hat{R}_{ts}^{Y}$, given the label, $Y_{ts}$, as predicted by the application of d-separation to the unconfounded model $C_{ts} \rightarrow Y_{ts} \rightarrow \hat{R}_{ts}$. (Note, as well, that while we also observe that the distribution of $cor(C, R)$ is centered at a value close to zero (indicating a weak marginal association between $\hat{R}_{ts}^{Y}$ and $C_{ts}$) we have that this is likely due to the weak association between $C_{ts}$ and $Y_{ts}$ observed in our data, and that, in general, deconfounded predictions might still show a marginal association between $C_{ts}$ and $\hat{R}_{ts}$ when both $C_{ts}$ and $Y_{ts}$ and $\hat{R}_{ts}$ and $Y_{ts}$ are strongly associated.) Analogous results for the PsA-without-UEI classifier are presented in Figure S18 b.

Finally, it is important to clarify that all the results presented here should be seen as preliminary given that our sample size is small, and results might change as we collect more data. Also, it is important the keep in mind that the confounding techniques described in this document are only applicable to observed confounders, and that our results might still be biased by unobserved confounders. Lastly, it is important to point out that the causality-aware adjustment should be preferred over performing a simple residualization

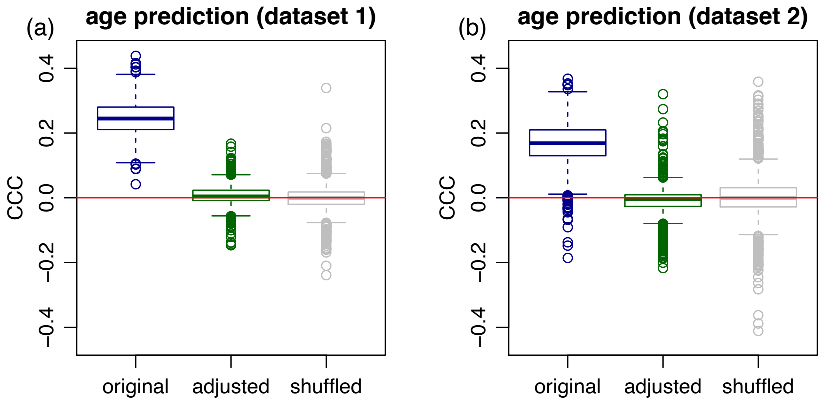


**Figure S17: Adjusted confounder predictions.** The figure compares the predictive performance of a regression model trained with the age-adjusted total-rotation (green boxplot) against a regression model trained with the unadjusted total-rotation (blue boxplot). As before, the grey boxplot report the results based on shuffled labels.

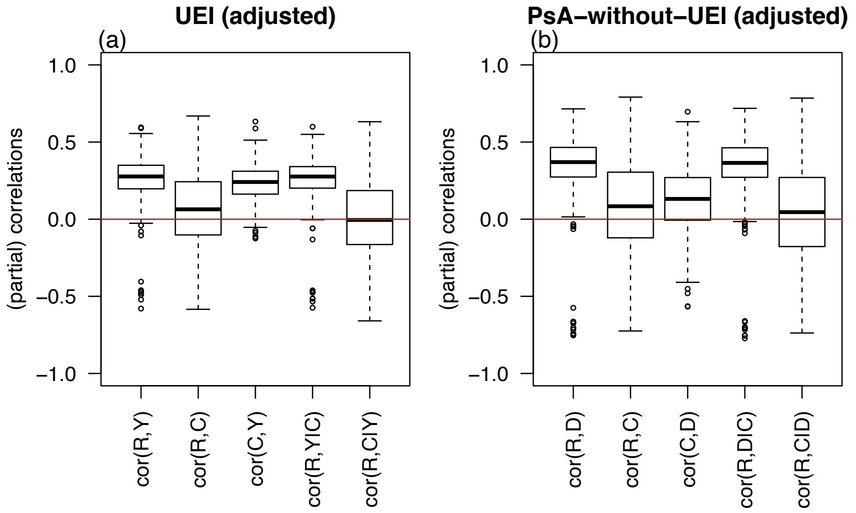


**Figure S18: CI pattern evaluations for classifiers trained with the adjusted total-rotation input.** $R$ represents the test set prediction $\hat{R}^Y_{ts}$ or $\hat{R}^D_{ts}$, $C$ represents the age, $Y$ represents the UEI label, and $D$ presents the disease status, namely, PsA (without UEI) or PsO.

adjustment (where we regress the inputs on the confounders alone, and use the residuals as the new inputs of the classifier). As described in [13], the residualization approach corresponds to the wrong adjustment in anticausal prediction tasks.

### Nail Object Detection and Nail Psoriasis Classification


**1. Overview** **1**
Figure 1: Workflow for nail psoriasis classification. 1
**2. Nail Object Detection** **2**
2.1 Dataset 2
2.2 Object Detection 2
Figure 2: Workflow for nail object detection using Detectron2 3
2.3 Model Fine-tuning 3
**3. Psoriasis Classification** **4**
3.1 Dataset 4
3.2 Model Architecture 4
**4. References** **5**


#### 1. Overview

Our workflow consisted of three main components: transfer learning, nail object detection and nail psoriasis classification on ‘cropped’ images of nails (Fig. 1). Transfer learning has been shown to be a powerful method for image analysis, particularly with smaller datasets. In our workflow, transfer learning was introduced into the model training by fine-tuning previously trained weights of Detectron2 Faster RCNN FPN model[1] on a customized, open source hand image dataset (see below) for nail tracking. Once the object detection model successfully detected nails, we used the VGG16 model[2] which was trained on the imagenet dataset and fine-tuned on cropped nail images to classify nail psoriasis vs not psoriasis.

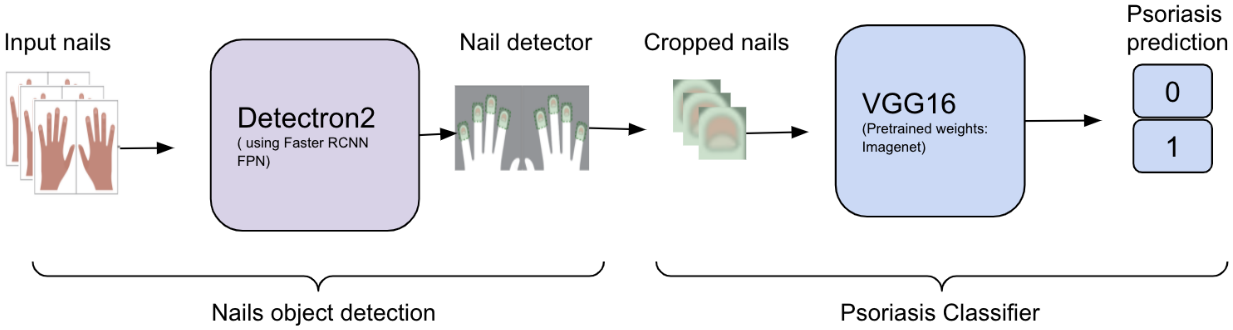


**Figure 1: Workflow for nail psoriasis classification.**

Fine-tuning of Detectron2 Faster RCNN and VGG16 pretrained models were used on nail images for the binary classification of nail psoriasis.

#### 2. Nail Object Detection

##### 2.1 Dataset

We derived a dataset for nail object detection using open source hand images from the Hong Kong Polytechnic University Contactless Hand Dorsal Images Database,[3] nail psoriasis images from the Pubmed Central database, and healthy nail images from a Google Image search. We aggregated 1020 image frames with a frame resolution of 416x416, where each image consists of at least one nail in it. After processing from these larger images of the hands, our dataset consisted of more than 5,000 individual nails segmented from all images.

The *LabelImg* annotation tool[4] was used to annotate bounding boxes across each nail from hand images. We also labeled the nail's section when partial nails blocked due to other fingers.This labeled dataset was distributed into training, validation and testing. The training set was used to update the weights of the model, validation was used to monitor the performance during the training phase and the test was the final hold out sample to evaluate the performance of the trained model.

##### 2.2 Object Detection

This subsection discusses our use of the Detectron2 Faster R-CNN,[1] a wrapper for object detection tasks with a deep neural network. We were interested in the detection of nails, to be followed by classification of psoriasis, and clinically it is possible that we might have psoriasis on the borders/edges of the nails. Thus to handle all the edge use cases, bounding box detection proves to be a much better solution compared to the segmentation in our case. Faster R-CNN is an easy framework for object detection and delivers best performance on COCO dataset, most commonly used for comparing object detection model performance.

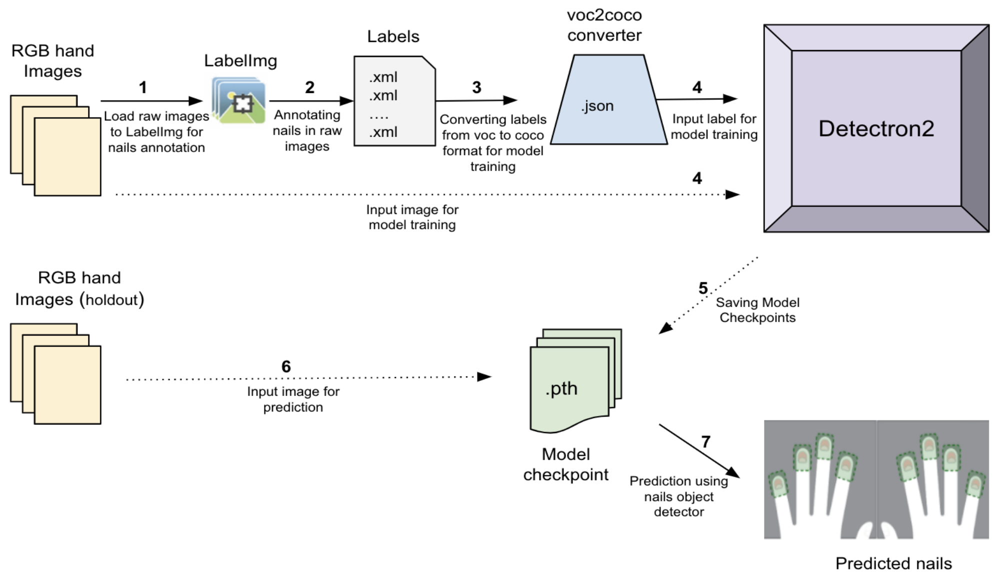


**Figure 2: Workflow for nail object detection using Detectron2**

#### 2.3 Model Fine-tuning

We fine-tuned the pretrained Faster R-CNN architecture on the training images. But in addition to images, Dectectron2 required images with their ground truth labels. As represented in Figure 2, to generate the ground truth annotations, first we needed to label the bounding box for all the nails in the hand images. We loaded all the raw images on *LabelImg* to annotate nails from a given image and save them in voc format. Our object detection algorithm expected input in coco format, so we converted all the labels from voc to coco format and fed images with their annotations into detectron2. During training, the optimizer tuned all the model parameters in a way that models generalized to corresponding ground truth. After training, the network received a new image from a holdout sample and plotted all the possible bounding box regions which had high probability for the presence of nails.

As a part of technical specification we used Pytorch 1.9.0 and trained the model with one NVIDIA GPU. For training, we used the default loss function, learning rate of 0.001 and batch size of 64 images, which achieved best performance on our validation sample. We trained the model for 1500 epochs and evaluated the performance on validation set after 500 epochs.

#### 2.4 Evaluation

We generated the two types of holdout dataset to evaluate the performance of the model, holdout set A (open source images from our dataset described above) and holdout set B (data collected and annotated in-clinic as a part of this study). Then, we used the mean Average Precision (mAP) score to compare the performance of the model, with a bounding box crossway from 50% to 95% intersection over union (IoU), where IoU is defined as area of intersection over area of union. For holdout set B, we also labeled the Fitzpatrick skin tone to evaluate model performance over different skin tones that represented how well fine-tuned model weights generalized across levels of pigmentation in our model.

### 3. Psoriasis Classification

#### 3.1 Dataset

For in-clinic, physician-labeled data, we had 316 images for controls and 38 images for nail psoriasis cases collected from the participants recruited from two medical centers , the New York University Langone Psoriatic Arthritis Center and the Brigham and Women's Hospital, between June 5, 2019 to November 10, 2021. To correct for the imbalance between nail psoriasis and healthy nails, we decided to use open source Pubmed Central nail psoriasis images (281 nail psoriasis images) to balance our sample. Our training samples for controls were from the study while our training samples from cases were completely from an open source dataset. Thus, our holdout case sample and training case samples were from different sources.

#### 3.2 Model Architecture

Model building and training was performed on Tensorflow version 2 (Google, Mountain View, California, USA). In our experiment, we used the VGG16 network, pre-trained on the imagenet dataset. For the transfer of feature vector from the tail layers and retraining of the model, we excluded the output layer of the network by include_top = False, and added a custom sequential layer which first flattened the feature vector then we added a dense layer of 256 fully connected units followed by a dropout. Then the final classifier output layer was added with two output units with softmax activation function. The input tensor to the model was 224x224x3 in dimension. To reduce the complexity and avoid overfitting, we froze all the layers of the VGG16 network except the last convolution layer just to make our network more generalizable on nail psoriasis data. To evaluate the performance of the model, 5 fold cross validation was used, however as we mentioned, case data in training sample was from open source dataset and not close enough to our study samples in terms of severity of psoriasis, so we could expect difference between validation and holdout sample recall score (only for case, as our training cases data was not from study). We used categorical cross entropy as a loss function, l2 loss regularization, keras stochastic gradient descent as optimizer with a learning rate of 0.0001 and training batch size was 20 images per batch.

##### 3.3 Evaluation

We reported the final results (accuracy, precision, recall etc.) on holdout samples that include unseen samples for cases. To further estimate the effect/variance of model performance on holdout samples, we conducted 1000 iterations of bootstrapping to calculate the 95% confidence interval for all the classification matrices.

2 Website. https://arxiv.org/abs/1409.1556.

3 The Hong Kong Polytechnic University Finger Image Database, Version 1.0. http://www4.comp.polyu.edu.hk/~csajaykr/knuckleV2.htm (accessed Jan 30, 2022).

4 Website. Tzutalin. LabelImg. Git code (2015). https://github.com/tzutalin/labelImg.

### Hand Imaging and Fitzpatrick Skin Tone

**Overview**

OpenCV[1], an openly available python library for computer vision applications, was used for all image and pixel manipulations. Mediapipe[2], an open cross platform framework for Machine learning pipelines, was used to detect hand landmarks. SuperAnnotate[3] was used to segment the hands from the background in a given image; this hand mask was used to estimate the contour of the hand. We then used the hand landmarks from mediapipe to guide the estimation of the Fitzpatrick skin tone, and in conjunction with the contour from SuperAnnotate to locate individual finger joints and estimate their thickness.

**Generating Hand Landmarks**

We used the hand and finger tracking solution of Mediapipe, Mediapipe-Hands, to estimate the hand landmarks for a single hand image. The landmarks estimated using are illustrated in Fig 1. Using OpenCV, we first resize the input hand image, in such a way that the maximum resolution of the height/width is 300 pixels (optimal for Mediapipe) while maintaining the aspect ratio. We then process the downsampled image through Mediapipe-Hands to get various hand landmarks. The position of these landmarks is relative to the dimensions of the image, as a ratio to the input height and width. We used the same landmarks to crop out relevant sections from the original image, by using the original image dimensions to recalculate the landmark location in actual pixels in the original image.

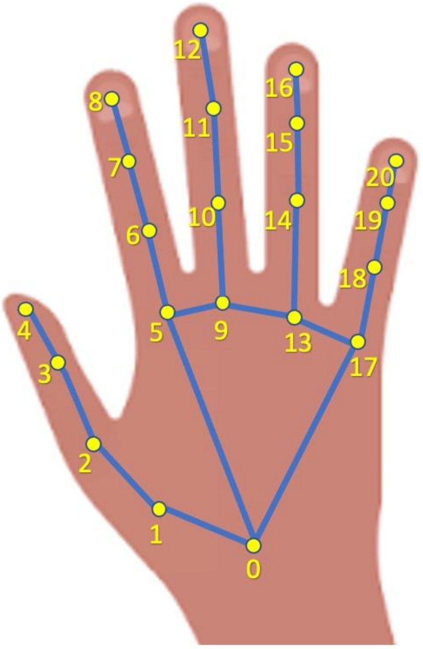


0. WRIST
1. THUMB_CMC
2. THUMB_MCP
3. THUMB_IP
4. THUMB_TIP
5. INDEX_FINGER_MCP
6. INDEX_FINGER_PIP
7. INDEX_FINGER_DIP
8. INDEX_FINGER_TIP
9. MIDDLE_FINGER_MCP
10. MIDDLE_FINGER_PIP
11. MIDDLE_FINGER_DIP
12. MIDDLE_FINGER_TIP
13. RING_FINGER_MCP
14. RING_FINGER_PIP
15. RING_FINGER_DIP
16. RING_FINGER_TIP
17. PINKY_MCP
18. PINKY_PIP
19. PINKY_DIP
20. PINKY_TIP

**FIGURE 1**

**Fitzpatrick Skin Tone estimation**

We approximate the skin tone of the individual from their hand by looking at the region formed by the boundary points corresponding to metacarpophalangeal (MCP) joints [5,9,13,17 from Fig 1] and the wrist [0 from Figure 1]. We cropped out this convex shape and converted each pixel from the RGB space to the CIEL*a*b space. We then used the L,b values from the CIEL*a*b space to calculate the Individual Typology Angle (ITA) for each pixel [Eqn 1]. ITA is a measure of constitutive pigmentation in the skin, as such we used it as a marker for the skin tone.

$$ITA = arctan((L - 50)/b) * 180/pi \quad \text{(Eqn 1)}$$

The ITA values from all pixels were summarized using a median, and the median ITA value was used to estimate the Fitzpatrick Skin Tone[4] using the thresholds given below.

| Fitzpatrick scale | ITA (Individual Typological Angle) |
|---|---|
| 1 | ITA > 55 |

| | |
|---|---|
| 2 | 41 < ITA <= 55 |
| 3 | 28 < ITA <= 41 |
| 4 | 10 < ITA <= 28 |
| 5 | -30 < ITA <= 10 |
| 6 | ITA < -30 |

**Estimation of Joint thickness**

Joint thickness is an indicator for swollen joints or dactylic fingers. We wanted to create a metric, an effective joint thickness measurement, which could be used to estimate if a joint was swollen. The first step was to crop out each finger from the hand image and then estimate the thickness of each individual joint in that finger.

We converted the input image into a black and white image, and segmented the hand using the mask obtained from SuperAnnotate. In the future we hope this segmentation process can be automated by an AI model. After getting the segmented hand mask, we used that to draw contours along the hand border. We then drew a convex hull that encloses the whole contour of the hand. We chose relevant landmarks obtained from Mediapipe-Hands as fingertip landmarks [8,12,16,20 from Fig 1]. We then calculated the points on the convex hull closest to the fingertip landmarks, and took that as a proxy for the actual tip of the finger. We did not consider the thumb in our pipeline, as we found it difficult to estimate the landmarks on the thumb. For many individuals, the thumb was facing the camera unlike all the other fingers, and sometimes it was out of view.

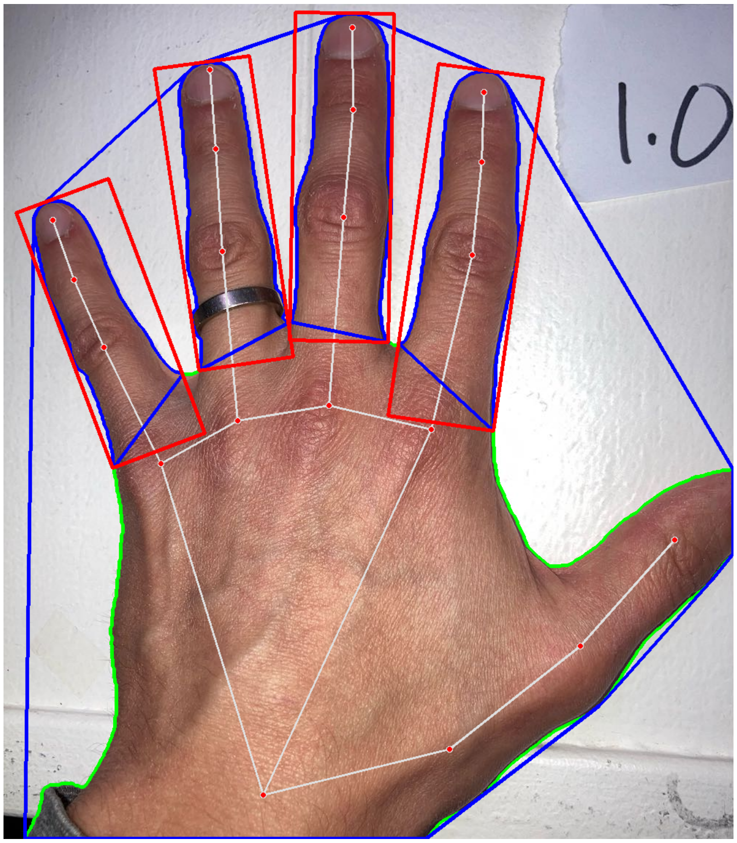


**Figure 2**

For each finger we interpret the contour in two ways, one to the left of the finger tip and another to the right. For each subsection of the contour, left and right, we found the point on that subsection that is closest to the corresponding MCP joint of the finger under consideration. From the main contour, we then subsetted the contour whose endpoints are the point on the convex hull corresponding to the fingertip, and the two closest points to
MCP joint on either side of this hull point to get the individual finger contour.

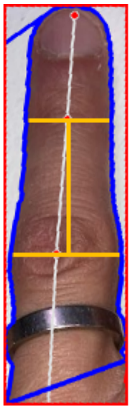

Once we had an individual finger contour, we used a minimum bounding rectangle around this contour to crop out the finger. A perspective transform (getPerspectiveTransform, warpPerspective in OpenCV) was used to orient the cropped finger images, so that the finger tip is pointing north. We used the landmarks from the original image to recalculate the new positions of the proximal interphalangeal (PIP) and distal interphalangeal (DIP) joints with respect to the individual cropped finger images. To calculate the width, we counted the number of pixels in the foreground in a row as a proxy for the width of the finger at the position corresponding to the row.

**Figure 3**

We calculate the pip to dip length of a given finger as the distance (number of pixels) between the two rows passing through the pip and dip joints. We calculate the effective width of a joint, as the ratio of the width of a joint to the average of pip to dip length of all fingers (except thumb)

Effective Width of PIP(DIP) = Width of PIP(DIP) / Mean pip to dip length across all fingers.
